# Tau PET Braak staging across the Alzheimer Disease continuum using Flortaucipir and MK6240

**DOI:** 10.64898/2026.11.01.26367034

**Authors:** Andreia Rocha, Carolina Soares, Marina Scop Medeiros, Emma Ruppert, Arthur C. Macedo, Firoza Z. Lussier, Guilherme Povala, Bruna Bellaver, Pamela C.L. Ferreira, Guilherme Bauer-Negrini, Livia Amaral, Matheus Rodrigues, Markley Oliveira-Junior, Rayan Mroue, Álvaro de Oliveira Franco, Cécile Tissot, João Pedro Ferrari-Souza, Nesrine Rahmouni, Jeffrey L. Evelhoch, Tobey J. Betthauser, Dana L. Tudorascu, Thomas K. Karikari, Joseph C. Masdeu, Juan Fortea, David N. Soleimani-Meigooni, Val Lowe, Hwamee Oh, Belen Pascual, Brian A. Gordon, Pedro Rosa-Neto, Suzanne Baker, Tharick A. Pascoal, HEAD Study

**Author notes:** Correspondence to Tharick A. Pascoal, MD, PhD. Department of Psychiatry and Neurology, University of Pittsburgh, Pittsburgh, PA, USA. 200 Meyran Ave - Parkvale Building, 5th Floor, Pittsburgh, PA, 15213, USA.

## Abstract

Tau positron emission tomography (PET) staging is increasingly used to stratify Alzheimer’s disease (AD), yet cross-tracer interchangeability remains unestablished. In a prospective head-to-head study of 682 individuals across the aging-to-AD continuum, we compared tau PET Braak staging derived from [^18^F]Flortaucipir and [^18^F]MK6240. While both tracers showed high internal adherence to hierarchical Braak patterns (89-92%), cross-tracer agreement was limited (43%, weighted κ = 0.54) and fell to 14- 23% at early-to-intermediate stages, a key window for therapeutic intervention. Discordance was strongly directional: in over 90% of discordant cases [^18^F]MK6240 assigned a higher stage, on average approximately one stage above [^18^F]Flortaucipir. [^18^F]MK6240 upstaging was associated with greater Aβ burden, adverse plasma biomarker profiles, and worse cognitive performance. Stage-frequency benchmarking against an external neuropathological cohort (N = 4,422) revealed that PET staging lags neuropathology by ∼2 Braak stages for [^18^F]MK6240 and ∼3 for [^18^F]Flortaucipir in the early phases of disease, with progressive convergence at later stages. Tau PET Braak staging is internally coherent within tracers but not interchangeable across them, with [^18^F]MK6240 systematically assigning higher stages than [^18^F]Flortaucipir. This biologically meaningful discordance, most pronounced at early stages, has direct implications for patient selection and staging for clinical trials and practice, necessitating tracer- aware harmonization strategies.

## Introduction

Alzheimer’s disease (AD) is neuropathologically characterized by the presence of amyloid-beta (Aβ) plaques and tau neurofibrillary tangles. The stereotyped spatial progression of tau lesions in AD brains led Braak and Braak to propose a six-stage framework, in which pathology emerges in transentorhinal regions, extends to limbic structures, and ultimately involves widespread neocortical areas(*1, 2*). This pathological progression typically unfolds over many years and, in most cases, follows a predictable spatial sequence, with increasing tau burden closely associated with clinical severity(*3–5*). This hierarchical staging framework subsequently became a central pillar of the neuropathological diagnosis of AD(*6–9*).

The advent of tau positron emission tomography (PET) tracers has enabled in vivo visualization and quantification of tau pathology. In this context, the Braak staging system has been adapted from histopathology to tau PET, to stratify living individuals based on their tau PET deposition patterns (*10–12*). PET-based Braak staging largely recapitulates the hierarchical tau progression observed at autopsy and correlates with clinical severity and disease biomarkers(*10–14*), but may systematically underestimate tau histopathology, particularly at early stages(*15–17*). The clinical relevance of tau PET staging is underscored by recent disease-modifying therapy trials, such as the donanemab studies, in which tau PET served as a selection and stratification tool to enrich for tau-positive participants and to evaluate treatment effects across tau severity stages, with emphasis on individuals with low-to-intermediate tau burden(*18, 19*).

However, tau PET tracers differ in binding properties and performance(*20*). [^18^F]Flortaucipir (Flortaucipir), the only U.S. Food and Drug Administration (FDA)-approved tau PET tracer (Tauvid), exhibits off-target binding in regions including the basal ganglia, choroid plexus and brainstem, as well as limited sensitivity in medial temporal regions(*20*). These properties constrain sensitivity for detecting tau pathology in vivo at early Braak stages, in contrast to postmortem studies showing a high frequency of medial temporal tau pathology in aging and preclinical AD(*7, 21–23*). [^18^F]MK6240 (MK6240), introduced more recently, offers greater dynamic range and reduced off-target signal in subcortical regions(*20, 24, 25*). However, MK6240 shows important off-target binding in the meninges and other non-parenchymal structures, which can cause spillover of signal into adjacent cortical regions and bias regional quantification (*26, 27*).

Although Flortaucipir and MK6240 are widely used in clinical and research settings, the degree to which they produce concordant in vivo tau stage classifications remains unclear, a question central to the reliable staging of AD (*28*). We recently demonstrated that tracer selection influences the frequency of tau detection across the aging and AD continuum, with MK6240 identifying more tau-positive individuals than Flortaucipir in both cognitively unimpaired and impaired groups(*29*). We now address a complementary question: whether these two tracers produce concordant Braak stage classifications within the same individuals. Braak stages were assigned independently for each tracer to quantify cross-tracer concordance, characterize the magnitude and stage dependence of discordance and identify clinical and biomarker factors associated with divergent classifications. In addition, we anchored our findings to an independent neuropathological cohort to contextualize in vivo stage distributions relative to postmortem Braak staging.

## Results

### Study Participants

We studied 644 middle-aged and older participants (69 ± 8.4 years old) imaged with Flortaucipir and MK6240, comprising 362 cognitively unimpaired (CU) individuals, 225 with mild cognitive impairment (MCI), and 57 with dementia (**Table 1**). CU individuals were younger than MCI and dementia participants, who did not differ from each other in age. Clinical groups differed in sex and *APOE* ε4 status, whereas education did not differ across groups. Aβ positivity and cognitive performance (Clinical Dementia Rating Sum of Boxes, CDR-SB; Mini-Mental State Examination, MMSE; Montreal Cognitive Assessment, MoCA) showed progressive worsening from CU to MCI to dementia. An additional group of 38 CU young adults (23 ± 2.0 years old) was used exclusively for tau PET Braak staging cut-off derivation (**Supplementary Table 1**). As an external cohort, data from the National Alzheimer’s Coordinating Center (NACC) were used to contextualize tau PET-based Braak staging frequencies against neuropathology. The NACC cohort comprised 4,422 participants (514 CU, 546 MCI, and 3,362 dementia; 80 ± 12 years old; **Supplementary Table 2**). Clinical groups differed across demographic and clinical characteristics, including sex and education, with increasing Aβ positivity and clinical severity (CDR-SB) from CU to MCI to dementia. Dementia participants were younger at death and had a shorter interval between last assessment and death, while postmortem interval did not differ across groups.

**Table 1.**
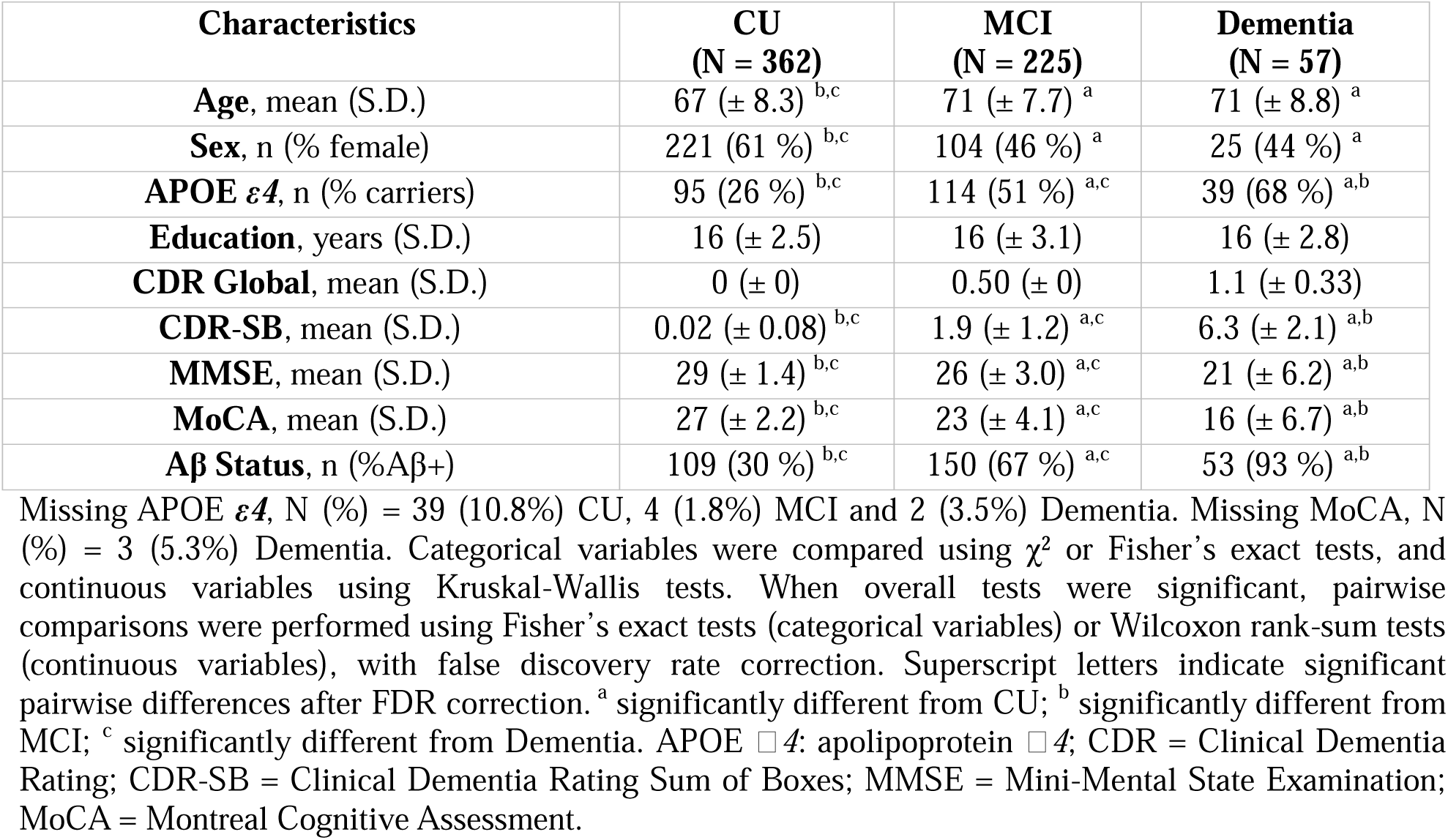
Demographics and key characteristics of HEAD Study participants.

| Characteristics | **CU<br>(N = 362)** | **MCI<br>(N = 225)** | **Dementia<br>(N = 57)** |
|---|---|---|---|
| **Age**, mean (S.D.) | 67 (± 8.3)<sup>b,c</sup> | 71 (± 7.7)<sup>a</sup> | 71 (± 8.8)<sup>a</sup> |
| **Sex**, n (% female) | 221 (61 %)<sup>b,c</sup> | 104 (46 %)<sup>a</sup> | 25 (44 %)<sup>a</sup> |
| **APOE *ε4***, n (% carriers) | 95 (26 %)<sup>b,c</sup> | 114 (51 %)<sup>a,c</sup> | 39 (68 %)<sup>a,b</sup> |
| **Education**, years (S.D.) | 16 (± 2.5) | 16 (± 3.1) | 16 (± 2.8) |
| **CDR Global**, mean (S.D.) | 0 (± 0) | 0.50 (± 0) | 1.1 (± 0.33) |
| **CDR-SB**, mean (S.D.) | 0.02 (± 0.08)<sup>b,c</sup> | 1.9 (± 1.2)<sup>a,c</sup> | 6.3 (± 2.1)<sup>a,b</sup> |
| **MMSE**, mean (S.D.) | 29 (± 1.4)<sup>b,c</sup> | 26 (± 3.0)<sup>a,c</sup> | 21 (± 6.2)<sup>a,b</sup> |
| **MoCA**, mean (S.D.) | 27 (± 2.2)<sup>b,c</sup> | 23 (± 4.1)<sup>a,c</sup> | 16 (± 6.7)<sup>a,b</sup> |
| **Aβ Status**, n (%Aβ+) | 109 (30 %)<sup>b,c</sup> | 150 (67 %)<sup>a,c</sup> | 53 (93 %)<sup>a,b</sup> |

Missing APOE ***ε4***, N (%) = 39 (10.8%) CU, 4 (1.8%) MCI and 2 (3.5%) Dementia. Missing MoCA, N (%) = 3 (5.3%) Dementia. Categorical variables were compared using χ<sup>2</sup> or Fisher’s exact tests, and continuous variables using Kruskal-Wallis tests. When overall tests were significant, pairwise comparisons were performed using Fisher’s exact tests (categorical variables) or Wilcoxon rank-sum tests (continuous variables), with false discovery rate correction. Superscript letters indicate significant pairwise differences after FDR correction. <sup>a</sup> significantly different from CU; <sup>b</sup> significantly different from MCI; <sup>c</sup> significantly different from Dementia. APOE □*4*: apolipoprotein □*4*; CDR = Clinical Dementia Rating; CDR-SB = Clinical Dementia Rating Sum of Boxes; MMSE = Mini-Mental State Examination; MoCA = Montreal Cognitive Assessment.

### Hierarchical Braak Staging Consistency in Flortaucipir and MK6240

To evaluate adherence to the hierarchical Braak scheme within each tracer, we quantified non- conformance, defined as positivity in a higher Braak stage region of interest (ROI) occurring in the absence of positivity in earlier Braak ROIs. Tau PET Braak staging was assessed using Flortaucipir and MK6240 under both the 7-class (0, I, II, III, IV, V, VI) and 4-class (0, I–II, III–IV, V–VI) schemes in the overall cohort (N = 644) and in the Aβ-positive subset (Aβ+, N = 312). Tau positivity for each tracer and ROI was defined as SUVR values exceeding the mean + 2 standard deviations (S.D.) of a young adult reference group.

Under the 7-class scheme, non-conformant classifications were infrequent and did not significantly differ between Flortaucipir and MK6240 in the overall cohort (9.3% vs 8.7%, *P* = 0.697; **Fig. 1a**) or in Aβ+ individuals (11% vs 8%; *P* = 0.225; **Fig. 1b**). Collapsing stages into the 4-class scheme further reduced non-conformity for both tracers in the overall cohort (3.6% vs 3.4%; *P* = 0.879; **Fig. 1a**) and in Aβ+ individuals (4.5% vs 2.6%; *P* = 0.193; **Fig. 1b**). The disruptions of the Braak hierarchy occurred more frequently at early and intermediate stages. For Flortaucipir (N = 60), the most common non-conformant pattern was Braak IV positivity in the absence of Braak II–III involvement, whereas for MK6240 (N = 56) the most frequent patterns involved Braak VI positivity without Braak V positivity or Braak III positivity without Braak I–II positivity (**Fig. 1c**). In Aβ+ individuals, these non-conformant patterns were largely consistent with those observed in the overall cohort.

**Figure 1.**
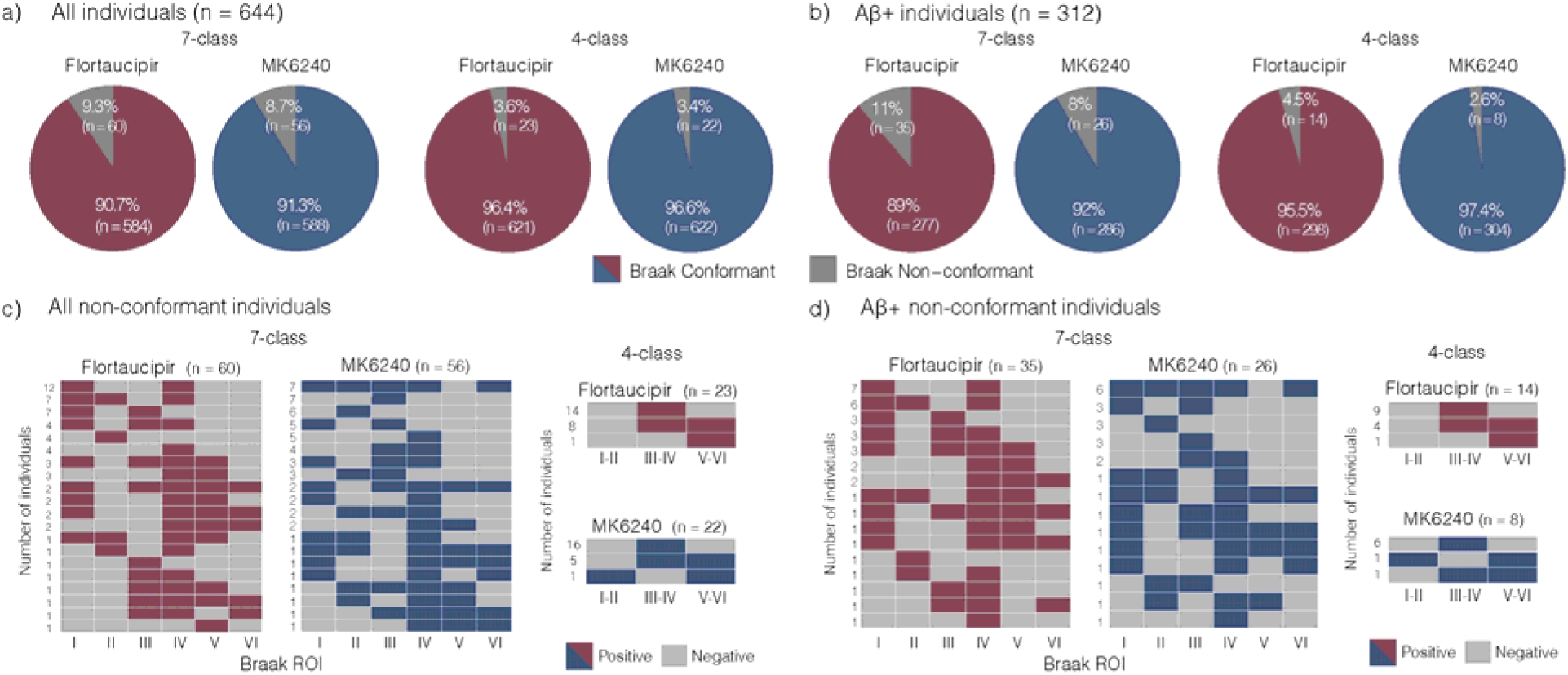
Flortaucipir and MK6240 follow the Braak hierarchy and non-conformant cases are tracer specific. **(a,b)** Proportion of individuals classified as Braak conformant or Braak non-conformant for Flortaucipir and MK6240 in the overall cohort (**a**, N = 644) and in Aβ+ participants (**b**, Centiloid ≥ 10; N = 312). **(c,d)** Binary matrices showing the distribution of Braak regional positivity patterns for Flortaucipir and MK6240 among individuals classified as Braak non-conformant in the overall cohort (**c**) and in Aβ+ participants (**d**). Binary matrices indicate positivity across Braak regions of interest (ROIs), with values representing the number of individuals exhibiting each non-conformant pattern. All results are presented under the 7-class and 4-class Braak staging schemes. Non-conformant cases represent individuals whose tau distribution did not follow the hierarchical Braak ordering.

Under the 4-class scheme for all analyses and groups, the predominant non-conformant pattern was Braak III–IV positivity in the absence of Braak I–II positivity (**Fig. 1d**). Of the 60 and 56 non- conformant cases for Flortaucipir and MK6240, respectively, only 14 involved the same individuals — and just 2 shared the same Braak ROI positivity pattern (**Supplementary Fig. 1**). Additionally, Braak stage distributions aligned with clinical severity and Aβ status for both tracers, with Aβ-negative (Aβ-) CU individuals predominantly classified as Braak 0 and progressively higher Braak stages observed in Aβ+ MCI and dementia (**Supplementary Fig. 2**). Using within-group comparisons between tracers we identified that differences in stage frequencies between Flortaucipir and MK6240 were confined to Aβ+ groups and limited to stages I and VI in the 7-class scheme and to stages 0 and V–VI in the 4-class scheme (**Supplementary Table 3**).

Voxel-wise analyses confirmed that both tracers demonstrated stepwise spatial progression from medial temporal to limbic and neocortical regions, consistent with Braak topography (**Fig. 2**). Similar regional patterns were observed in the frequency of abnormal voxels and Z-scored brain maps (**Supplementary Fig. 3**).

**Figure 2.**
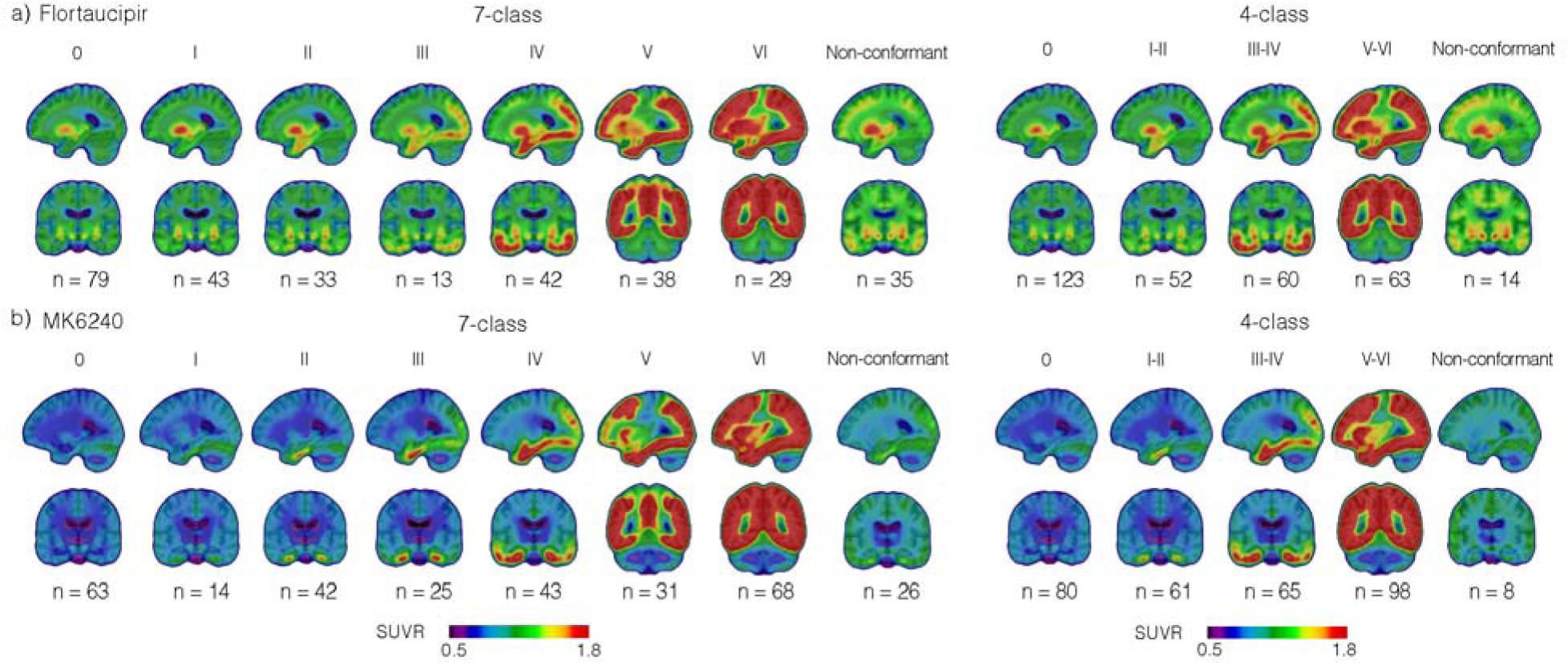
Voxel-wise Flortaucipir and MK6240 PET Braak stages. Group-averaged tau PET images stratified by Braak stage for Flortaucipir (**a**) and MK6240 (**b**). Representative sagittal and coronal slices are shown. All results are presented under the 7-class and 4- class Braak staging schemes. All analyses were restricted to Aβ+ individuals (Centiloid ≥ 10; N = 312). Non-conformant cases represent individuals whose tau distribution did not follow the hierarchical Braak ordering and are shown separately.

### Agreement between Flortaucipir- and MK6240-based Braak staging

We next evaluated agreement in Braak stage assignment between Flortaucipir and MK6240 in Aβ+ individuals, restricting subsequent analyses to this subset to focus on AD pathology. Under the 7-class scheme, less than half of individuals (43%, weighted κ = 0.54; **Fig. 3a,b**) showed concordant Braak stage assignments between Flortaucipir and MK6240. Collapsing stages into the 4-class scheme increased concordance to 57% (weighted κ = 0.59, **Fig. 3d,e**). Results stratified by clinical groups can be found in **Supplementary Fig. 4**. Concordance analyses using alternative positivity cut-offs and mixed combinations yielded similar discordance rates, with only minor variations largely driven by differences in Braak 0 or non-conformant classifications (**Supplementary Fig. 5**).

**Figure 3.**
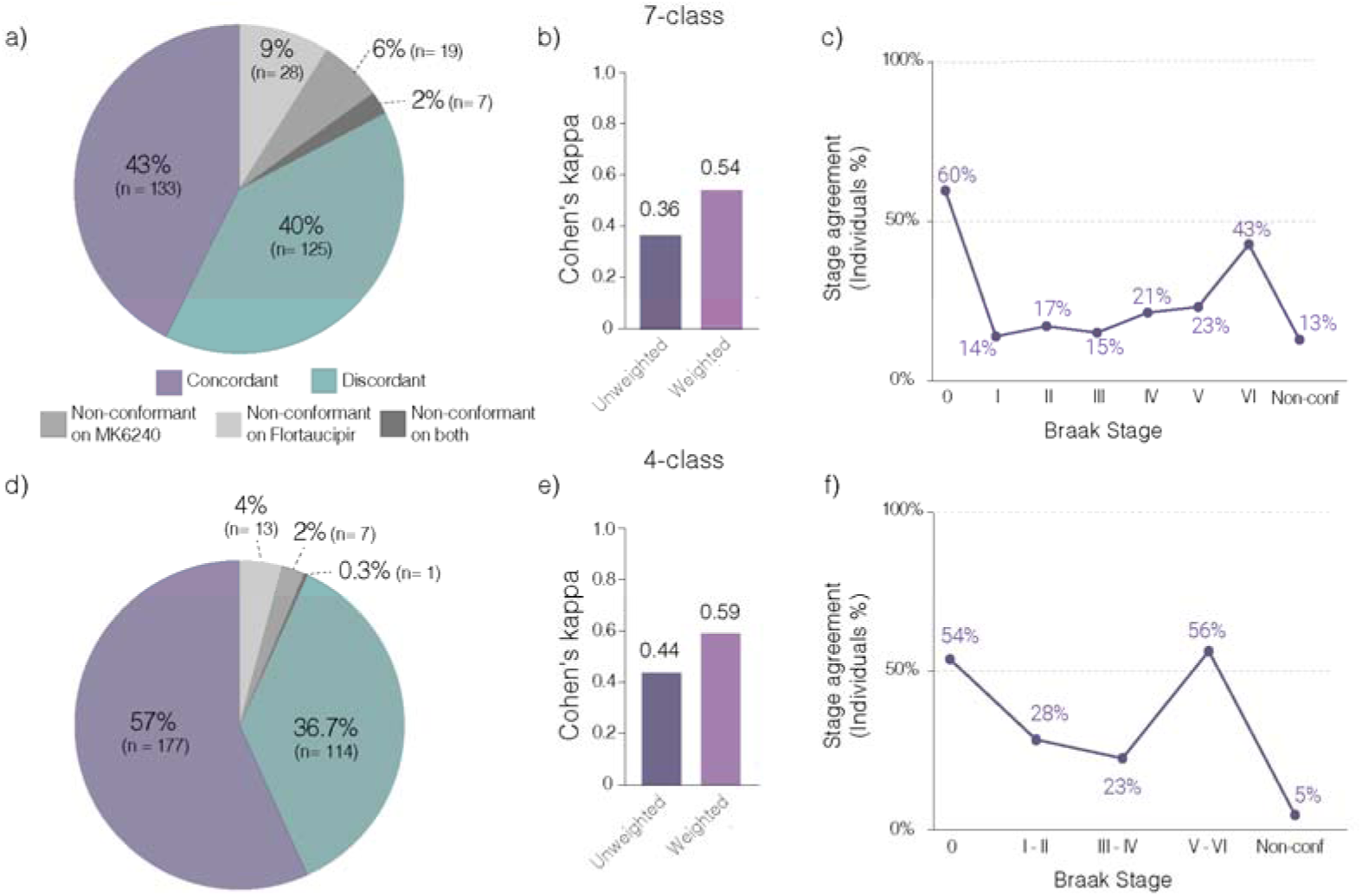
Flortaucipir and MK6240 show moderate agreement in Braak stage assignment. **(a,d)** Proportion of individuals classified as Braak concordant, Braak discordant, or Braak non- conformant between Flortaucipir and MK6240 under the 7-class (**a**) and 4-class (**d**) Braak staging schemes. **(b,e)** Inter-tracer agreement quantified using unweighted and weighted (linear) Cohen’s κ statistics for the 7-class (**b**) and 4-class (**e**) Braak staging schemes. **(c,f)** Agreement by Braak stage, showing the proportion of individuals assigned to the same Braak stage by both tracers under the 7-class (**c**) and 4-class (**f**) Braak staging schemes. Concordant = identical Braak stage assigned by both tracers; discordant = different Braak stage assigned by the tracers; non-conformant = hierarchical Braak ordering not followed by one or both tracers. All analyses were restricted to Aβ+ individuals (Centiloid ≥ 10; N = 312).

Under the 7-class scheme, agreement was highest at Braak 0 (60%) and Braak VI (43%), but markedly lower at early-intermediate stages: 14% at Braak I, 17% at Braak II, 15% at Braak III, 21% at Braak IV, and 23% at Braak V (**Fig. 3c**). Under the 4-class scheme, concordance peaked at Braak 0 (54%) and Braak V–VI (56%), with the lowest agreements at Braak I–II (28%) and at Braak III–IV (23%; **Fig. 3f**).

We next examined the direction and magnitude of inter-tracer differences in Braak stage assignment among discordant individuals. Under the 7-class scheme, discordant cases predominantly reflected higher MK6240-assigned stages (90.4%, 113 cases), typically differing by one or two stages, whereas higher Flortaucipir-assigned cases were rare (9.6%, 12 cases; **Fig. 4a,b**). A similar pattern was observed under the 4-class scheme, with most discordant individuals again assigned higher stages with MK6240 (87.7%, 100 cases) compared with Flortaucipir (12.3%, 14 cases; **Fig. 4c,d**).

**Figure 4.**
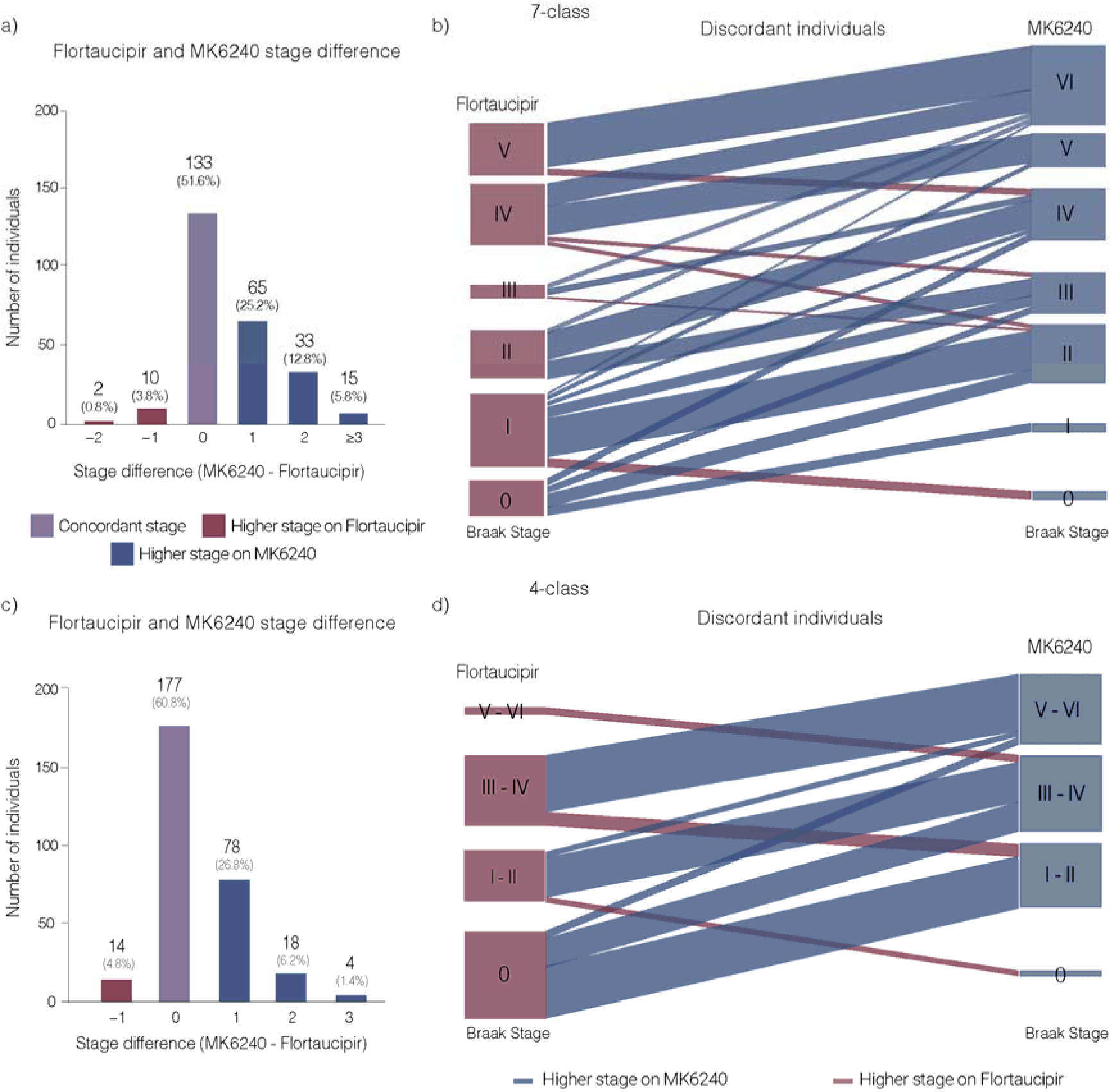
Individual-level inter-tracer Braak stage differences reveal a systematic directional bias favoring higher MK6240 assignments. **(a,c)** Distribution of inter-tracer Braak stage differences computed as MK6240 minus Flortaucipir. Results are shown for the 7-class (**a**, N = 258) and 4-class (**c**, N = 291) Braak staging schemes. Positive values indicate higher Braak stage assignment by MK6240, whereas negative values indicate higher stage assignment by Flortaucipir. **(b,d)** Alluvial plots illustrating transitions between MK6240- and Flortaucipir-assigned Braak stages for discordant individuals under the 7-class (**b**, N = 125) and 4-class (**d**, N = 114) Braak staging schemes. Flow widths represent the number of individuals. All analyses were restricted to Aβ+ individuals (Centiloid ≥ 10) and excluded Braak non-conformant cases. Discordant individuals are those that were assigned different stages by the tracers.

To facilitate cross-tracer comparisons, we developed probability-based harmonization matrices using the observed stage pairings between Flortaucipir and MK6240 (**Supplementary Fig. 6**). Under the 7-class scheme, several intermediate stages showed substantial overlap across adjacent categories rather than a unique one-to-one correspondence. For example, MK6240 Braak II corresponded to both Flortaucipir Braak I (45.9%) and II (29.7%), whereas Flortaucipir Braak II corresponded similarly to MK6240 Braak II (34.4%), III (25%) and IV (37.5%). Likewise, MK6240 Braak V showed comparable correspondence with Flortaucipir Braak IV (46.4%) and V (46.4%). Collapsing stages into the 4-class framework reduced this ambiguity and resulted in stronger correspondence between tracers. These harmonization matrices provide a probabilistic framework for translating Braak stage assignments between Flortaucipir and MK6240 and highlight that tracer-specific stage classifications cannot be assumed to be directly interchangeable.

To examine the relationship between continuous SUVR values and stage assignments, we plotted Flortaucipir SUVR as a function of MK6240-assigned Braak stage and compared it with values from the young reference group (**Fig. 5a, c**). Across Braak ROIs, most MK6240-positive individuals had Flortaucipir SUVR values overlapping with, or close to, the young reference group mean and well below the positivity threshold (46% to 56% of cases). These findings suggest that lowering the Flortaucipir threshold sufficiently to reclassify these cases as positive would likely also result in the misclassification of young healthy controls as tau positive. In the reciprocal analysis (**Fig. 5b,d**), plotting MK6240 SUVR as a function of Flortaucipir-assigned Braak stage, most Flortaucipir-positive individuals showed MK6240 SUVR values above the positivity threshold, with only occasional values approaching or falling below the reference threshold (6% to 8% of cases).

**Figure 5.**
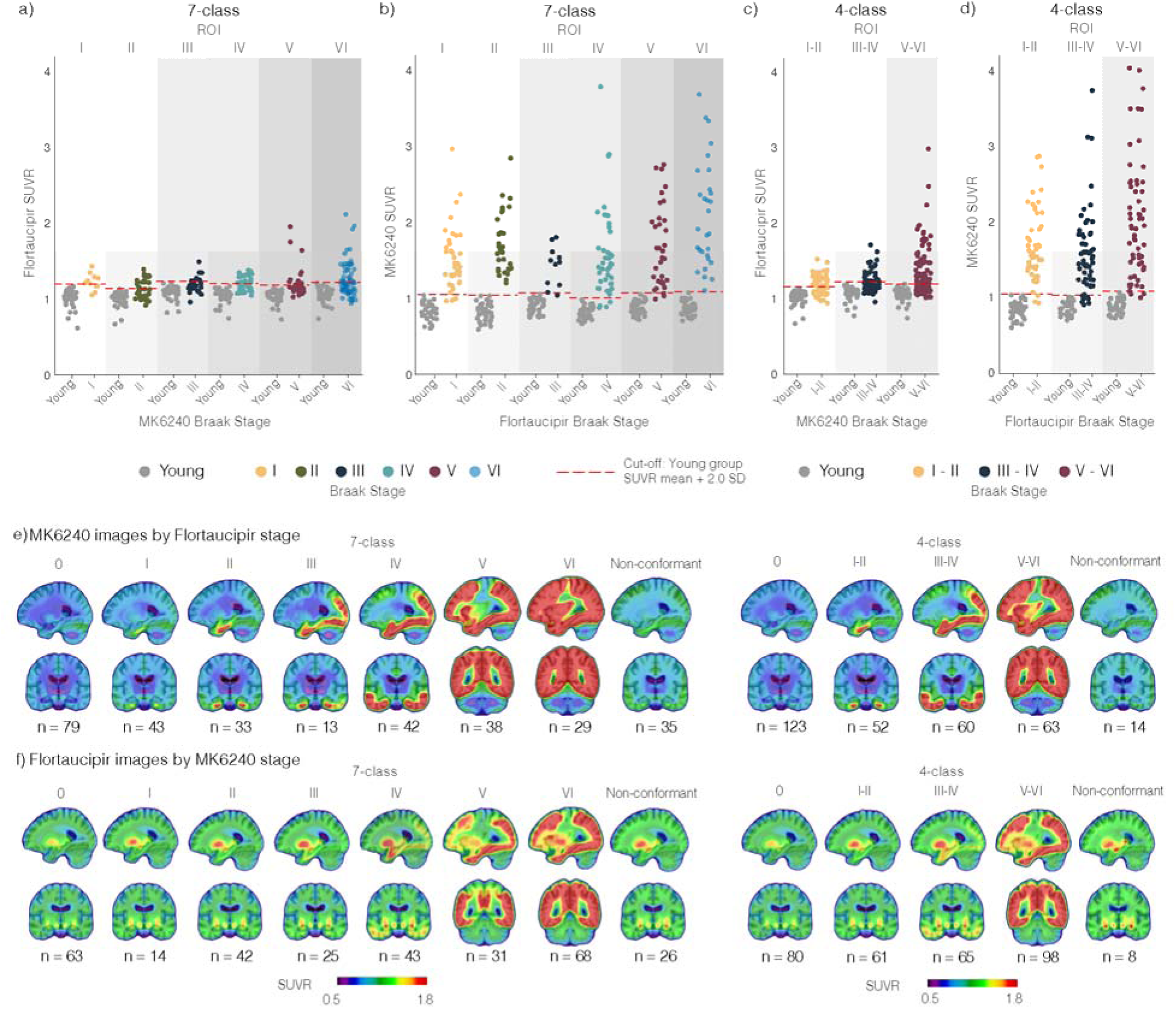
Cross-tracer comparisons reveal MK6240 captures tau pathology undetected by Flortaucipir at matched Braak stages. **(a, c)** Flortaucipir and **(b, d)** MK6240 regional SUVR values plotted as a function of the opposite tracer’s assigned Braak stage across Braak ROIs. Young individuals were plotted as reference group for each Braak ROI. Horizontal dashed line indicates positivity cut-off derived from young reference group mean + 2.0 S.D. **(a-d)** Non-conformant and Braak 0 individuals were excluded from this analysis (**a**, N = 223; **b**, N = 198; **c**, N = 224; **d**, N = 175; Young, N = 38). **(e)** Group-averaged MK6240 images stratified by Braak stage as assigned by Flortaucipir. Images illustrate the spatial distribution of MK6240 uptake corresponding to Flortaucipir-based Braak stage classification. **(f)** Group-averaged Flortaucipir images stratified by Braak stage as assigned by MK6240. Images illustrate the spatial distribution of Flortaucipir uptake corresponding to MK6240-based Braak stage classification. For both **(e,f)** panels, representative sagittal and coronal slices are shown and non-conformant cases, which represent individuals whose tau distribution did not follow the hierarchical Braak ordering, are shown separately. All analyses were restricted to Aβ+ individuals. All results are presented under the 7-class and 4-class Braak staging schemes.

To further assess differences in the spatial patterns of tracer uptake, we stratified individuals by one tracer’s Braak stage assignment and generated group-averaged images for the other tracer (**Fig. 5e-f).** When individuals were grouped by their Flortaucipir-assigned stage, group-averaged MK6240 images showed uptake extending beyond the regions that defined the Flortaucipir stage assignment; for example, Flortaucipir Braak I cases exhibited MK6240 signal extending into Braak II regions (**Fig. 5e**). Conversely, when grouped by MK6240-assigned stage, averaged Flortaucipir images showed less extensive spatial involvement, with uptake patterns corresponding to earlier Braak stages (**Fig. 5f**). These systematic differences in spatial extent at matched nominal stages were consistent across both staging schemes. Similar patterns were observed in the frequency of abnormal voxels and Z-scored brain maps (**Supplementary Fig. 7**). Furthermore, to quantify these spatial differences in spatial extent, we compared the proportion of abnormal voxels across Braak ROIs between tracers. Across Flortaucipir-defined Braak stages, the difference in abnormal voxel percentage (Δ = MK6240 − Flortaucipir) was predominantly positive across ROIs, indicating greater regional involvement detected by MK6240 (**Supplementary Fig. 8**). This difference peaked early: among Flortaucipir Braak I individuals, MK6240 identified 33.0% more abnormal voxels within Braak I and 21.1% more within Braak II, with 77.0% and 82.0% of participants, respectively, showing greater abnormal spatial extent with MK6240. At advanced stages, the effect became near-universal at the individual level, with 93–100% of Braak VI participants showing greater MK6240 spatial extent across Braak III–VI ROIs, a pattern confirmed under the 4-class scheme (8.3% difference, 90.3% of participants at stage V–VI). Representative cases are illustrated in **Supplementary Fig. 9**.

### Biomarker and clinical differences in MK6240-upstaged individuals

We next characterized individuals assigned to a higher Braak stage by MK6240 relative to Flortaucipir under both the 7-class and 4-class staging schemes (**Supplementary Fig. 10; Fig. 6; Supplementary Tables 6 and 7**). The clearest group differences were observed in the 4-class scheme, which provided greater statistical power by combining adjacent Braak stages (**Fig. 6**). Individuals assigned to a higher stage by MK6240 showed systematically greater biological and clinical severity than those assigned the same stage by both tracers. Among subjects classified as Braak 0 by Flortaucipir, MK6240-upstaged individuals exhibited higher Aβ burden (Centiloid; 64.3 ± 40.0 vs 38.5 ± 34.2, P = 0.0005), elevated plasma p-tau217 (0.78 ± 0.40 vs 0.46 ± 0.29 pg/mL, P < 0.0001) and GFAP levels (163.4 ± 83.0 vs 112.6 ± 54.6 pg/mL, P = 0.0009), higher CDR-SB scores (1.25 ± 1.59 vs 0.63 ± 1.38, P = 0.032), and lower MMSE (26.7 ± 3.1 vs 28.5 ± 1.9, P = 0.0006) and MoCA scores (23.5 ± 4.9 vs 26.1 ± 3.7, P = 0.003). Within Braak I–II, plasma p-tau217 remained significantly different between groups (1.07 ± 0.41 vs 0.75 ± 0.22, P = 0.0017), whereas no other measures differed significantly. No significant differences were observed at Braak III–IV. Analyses using the 7-class scheme yielded a similar overall pattern (**Supplementary Fig. 10**), with significant differences primarily observed at Braak stages 0 and I, although fewer comparisons reached statistical significance owing to the smaller sample sizes within individual Braak stages.

**Figure 6.**
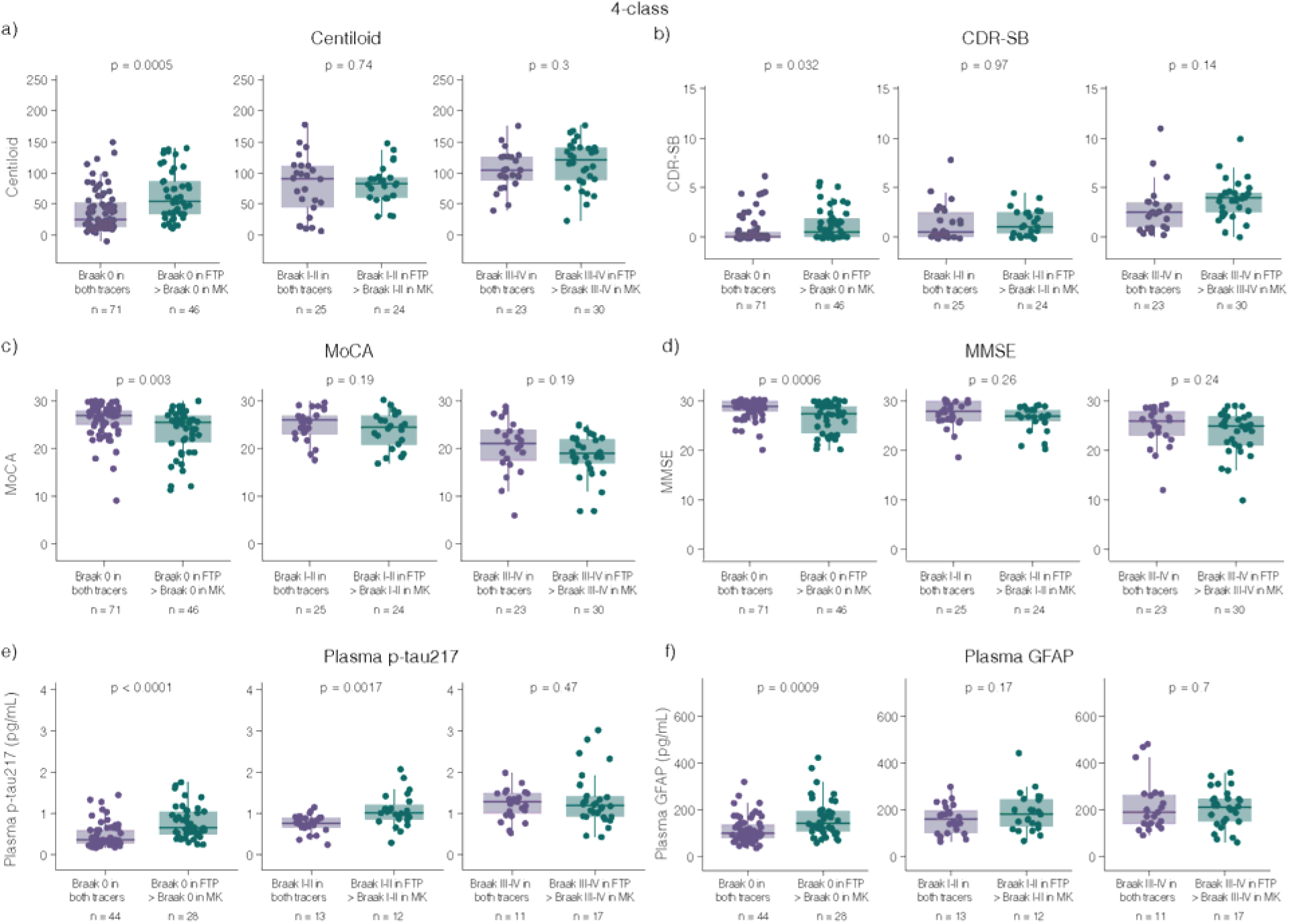
Higher MK6240-based Braak staging identifies individuals with greater biological and cognitive severity. Distributions of Aβ burden (Centiloid; **a**), cognitive performance (CDR-SB, MoCA and MMSE; **b**, **c, d**), and plasma biomarkers (p-tau217 and GFAP; **e**, **f**) in Aβ+ individuals grouped by Flortaucipir-based Braak stage and further stratified by whether MK6240 assigned the same stage or a stage higher classification under the 4-class Braak staging scheme. All analyses were restricted to Aβ+ individuals (Centiloid ≥ 10). Points represent individual participants. Boxes indicate the interquartile range (25th– 75th percentile), the central line represents the median, and whiskers extend to 1.5 × the interquartile range. Individuals classified as Braak non-conformant and those assigned to a higher Braak stage by Flortaucipir than MK6240 were excluded from this analysis (N = 277). FTP = Flortaucipir; MK = MK6240. CDR-SB = Clinical Dementia Rating Sum of Boxes; MMSE = Mini-Mental State Examination; MoCA = Montreal Cognitive Assessment.

### Tau PET Braak stage frequencies in relation to neuropathology frequency

We compared tau PET-derived frequencies of Braak ROI positivity in the HEAD Study with neuropathological Braak ROI positivity frequencies from an external autopsy cohort (NACC), stratifying individuals by cognitive performance (CDR-SB; **Fig. 7; Supplementary Tables 8 and 9**). Neuropathology consistently showed more frequent tau involvement than either PET tracer at lower CDR-SB levels, with PET-based regional positivity progressively approaching neuropathology frequencies as cognitive impairment increased.

**Figure 7.**
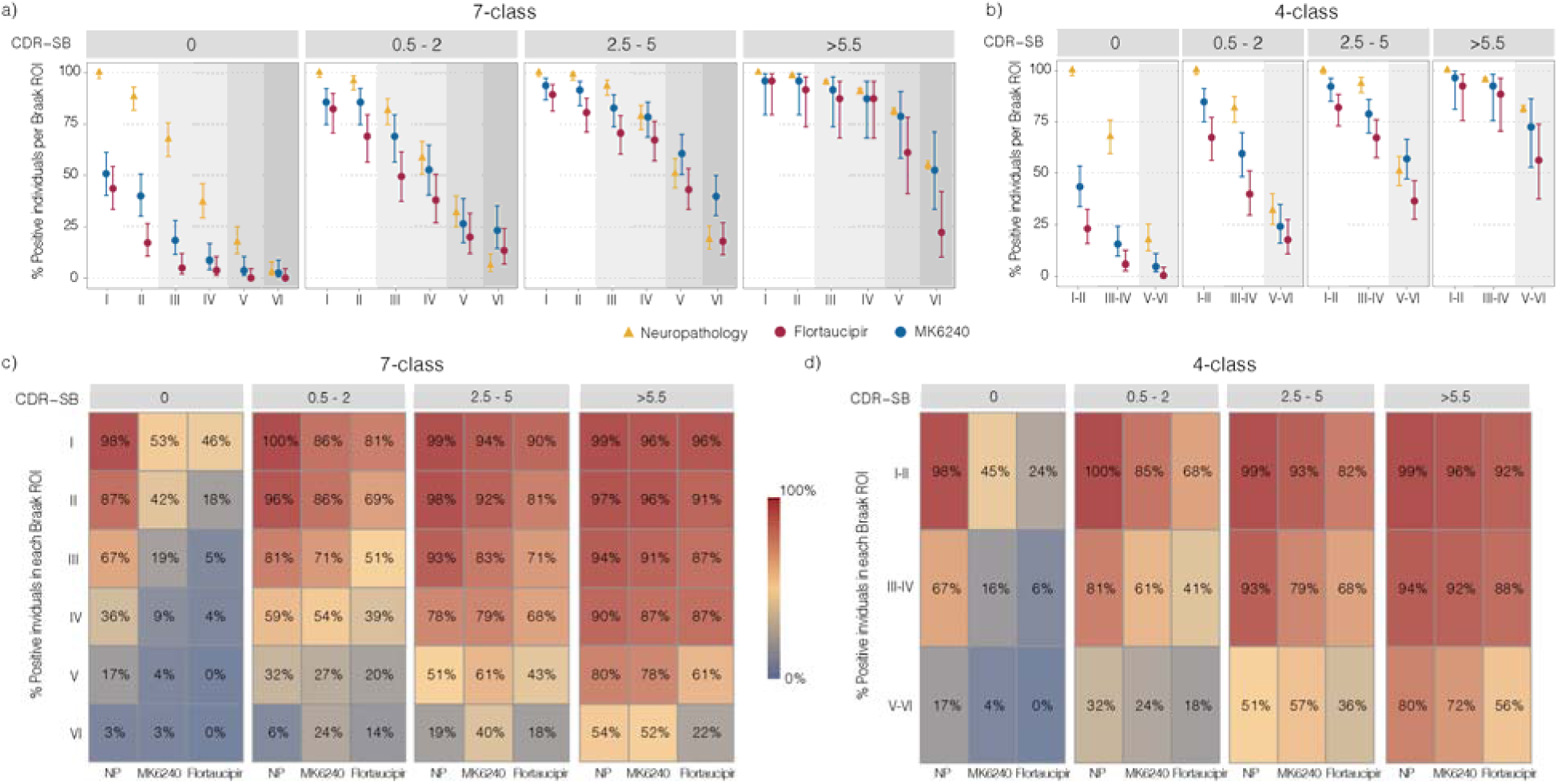
Mismatch between tau PET and neuropathological Braak stage frequencies across the AD spectrum. Dot plots and heat maps representing the proportion of tau-positive individuals in each Braak ROI stratified by cognitive performance using CDR-SB categories under the 7-class (**a, c**) and 4-class (**b, d**) Braak staging schemes. Neuropathology data are derived from the NACC cohort and PET data from the HEAD Study cohort. Dots represent group-level proportions and error bars the 95% confidence interval. All analyses were restricted to Aβ+ individuals (CERAD 2/3 for neuropathology or Centiloid ≥ 10 for PET) and excluded Braak non-conformant cases. Sample sizes per CDR-SB category: Neuropathology 0 (N = 129), 0.5–2 (N = 145), 2.5–5 (N = 182), >5.5 (N = 2286); PET 7-class 0 (N = 79), 0.5–2 (N = 59), 2.5–5 (N = 90), >5.5 (N = 23); PET 4-class 0 (N = 89), 0.5–2 (N = 74), 2.5–5 (N = 96), >5.5 (N = 25). NP = neuropathology.

These differences were statistically significant across early Braak stages in CU Aβ+ individuals (CDR-SB = 0) for both tracers after false discovery rate (FDR) correction, with odds ratios consistently <1, indicating lower PET-detected positivity relative to neuropathology. At CDR-SB 0, the Braak IV ROI positivity frequency observed with neuropathology corresponded to the postmortem frequency of Braak stages II and I in MK6240 and Flortaucipir respectively, reflecting a systematic shift of ∼2 and ∼3 stages lower than neuropathology.

As cognitive impairment increased (CDR-SB 0.5–2 and 2.5–5), PET-based positivity frequencies rose across Braak regions for both tracers and statistical differences diminished, with stage correspondence improving to roughly 1 stage below neuropathology or overlapping in the moderate impairment range. At the highest CDR-SB levels (>5.5), most stage-specific differences were no longer significant. A sensitivity analysis using 1:1 matched samples based on age, sex, and CDR-SB (**Supplementary Fig. 11**) showed similar results. To further facilitate cross-platform interpretation, we derived probability-based conversion tables mapping PET-based Braak stages to their most likely neuropathological equivalents, stratified by cognitive severity (**Supplementary Fig. 12**).

## Discussion

In this large head-to-head comparison of the two most widely used tau PET tracers, Flortaucipir and MK6240, across the AD continuum, PET-based Braak staging was hierarchically robust at the group level but highly tracer-dependent at the individual level. Inter-tracer agreement in Braak staging was only moderate, with more pronounced discordance in early and intermediate stages. Discordant classifications exhibited a directional asymmetry, with MK6240 consistently assigning higher Braak stages than Flortaucipir in most cases and, accompanied by a broader spatial extent of detected tau pathology and greater biological and clinical severity among upstaged individuals.. These differences could not be resolved using multiple techniques for cut-off determination. These findings demonstrate that tracer choice can substantially influence inferred tau stage, with direct implications for the interpretation, comparison, and harmonization of tau PET-based Braak staging across research studies and clinical settings.

In our study, both Flortaucipir and MK6240 demonstrated strong adherence to the expected Braak hierarchy and topographical progression. Hierarchical non-conformity was infrequent, particularly under the collapsed 4-class staging scheme. These observations are consistent with prior tau PET studies showing that in vivo imaging reliably recapitulates the hierarchical topographical progression of neurofibrillary pathology described in postmortem studies(*10, 11, 30–33*). When non-conformity occurred, uptake patterns most often involved regions susceptible to off-target binding or spillover effects for each tracer, such as basal ganglia and meninges, although some cases likely represent atypical distributions of tau pathology that have previously been described in AD(*34*). The limited overlap in non- conformant individuals between tracers further suggests that these deviations primarily reflect tracer- specific imaging characteristics rather than systematic deviances from the Braak framework itself. Together, these findings support the hierarchical conformance of PET-based Braak staging with the postmortem neuropathological stages originally proposed by Braak and Braak(*1*), regardless of the tracer employed.

Although each tracer demonstrated robust internal conformance with the Braak staging framework, inter-tracer agreement was only moderate, with concordance rates of 43% for the 7-class scheme and 57% for the 4-class scheme. Importantly, agreement was stage-dependent rather than uniform across the disease spectrum. Concordance was highest at Braak stage 0 and advanced neocortical stages (VI), whereas early and intermediate stages (I–V) – corresponding to periods of active tau propagation – showed the lowest agreement. Analysis of inter-tracer stage differences further revealed a pronounced directional asymmetry. In most discordant cases (90.4%), MK6240 assigned a higher Braak stage than Flortaucipir, typically by one or two stages, whereas instances in which Flortaucipir assigned a higher stage were rare (9.6%).

Further voxel-wise analyses and cross-tracer group-averaged images demonstrated that MK6240 systematically detected a broader spatial extent of tau pathology than Flortaucipir. When individuals were grouped according to their Flortaucipir-defined Braak stage, group-averaged MK6240 images consistently showed tau uptake extending beyond the regions defining the corresponding Flortaucipir stage. Conversely, when individuals were grouped according to MK6240-defined Braak stage, Flortaucipir images exhibited more spatially restricted uptake, frequently corresponding to an earlier stage of Braak progression. These differences were further quantified by abnormal voxel analyses, which demonstrated predominantly positive differences in abnormal voxel percentages across corresponding Braak regions. The largest differences were observed during the earliest stages of disease, with MK6240 detecting up to 43% more abnormal voxels in early Braak regions, and with the majority of individuals exhibiting greater abnormal voxel involvement with MK6240 in all stages. Together, these findings indicate that the higher Braak stages assigned by MK6240 reflect a broader spatial extent of detected tau pathology.

Importantly, individuals upstaged by MK6240 also showed higher Aβ burden, elevated plasma p- tau217 and GFAP levels, and worse cognitive performance compared to those assigned concordant stages, indicating that the additional tau signal captured by MK6240 reflects measurable biological and clinical disease. Most statistically significant differences were observed among individuals classified as Braak 0 by Flortaucipir, whereas fewer comparisons reached significance at later stages and under the more granular 7-class framework. Given the substantially smaller sample sizes within these stage-specific subgroups, the reduced number of significant findings likely reflects lower statistical power rather than the absence of biological differences. Consistent with this interpretation, effect sizes were generally comparable across the 4-class and 7-class analyses, despite the lower frequency of statistically significant comparisons in the 7-class framework.

The observed discordance was also robust across multiple thresholding strategies. Applying alternative positivity cut-off strategies, including mixed cut-off combinations applied differentially to each tracer, resulted in only minor changes in inter-tracer concordance. Rather than improving agreement, these approaches primarily altered the number of individuals classified as Braak 0 or as non-conformant, without substantially affecting concordance across the remaining Braak stages. Moreover, our analyses demonstrated that achieving concordance would require unrealistically low Flortaucipir cut-offs that would also classify a substantial proportion of young Aβ- individuals as tau-positive, undermining tracer specificity. The reverse adjustment would also carry a symmetric cost: raising MK-6240 thresholds to match Flortaucipir would exclude clearly positive individuals and compromise sensitivity. These findings indicate that inter-tracer discordance likely reflects intrinsic differences in tracer uptake characteristics and sensitivity to regional tau pathology, rather than cut-off-dependent artifacts.

This supports the notion that the directional asymmetry in stage assignment between MK6240 and Flortaucipir likely reflects fundamental pharmacological differences between the tracers(*20, 35–37*). MK6240 exhibits higher binding affinity for tau aggregates (Kd = 0.15–0.32 nM)(*35*) compared with Flortaucipir (Kd ranging from 0.57 to 14.6 nM depending on assay conditions)(*38, 39*), a difference that becomes functionally significant at the low tau densities characteristic of early Braak stages. Moreover, head-to-head autoradiography demonstrated that MK6240 has a wider effective dynamic range, driven by both higher specific binding in AD tissue and lower non-specific uptake in control tissue(*36*). These properties are further complemented by a more favorable off-target binding profile: Flortaucipir’s prominent choroid plexus signal spills into adjacent medial temporal regions, raising the baseline noise floor, whereas MK6240’s off-target binding is predominantly meningeal/skull-based and more spatially separable from Braak regions(*37, 40*). Collectively, these differences provide a mechanistic framework for understanding why MK6240 may capture tau signal earlier in the disease course, consistent with prior work showing MK6240 detects transentorhinal tau in 68% of Aβ-positive cognitively unimpaired individuals versus ∼20% with first-generation tracers(*11*).

Finally, to assess whether the frequency of tau PET-defined Braak stages aligned with neuropathological Braak stage frequencies, we anchored PET-based Braak staging to neuropathology using CDR-SB as a common measure of cognitive severity across the HEAD PET cohort and the NACC autopsy cohort. This strategy allowed the comparison of tau burden frequencies at equivalent levels of clinical impairment. Although indirect and imperfect, given that the neuropathology and tau PET data were not derived from the same individuals, this approach provides an informative framework to estimate the extent to which tau PET staging lags behind neuropathological staging. Comparisons inferred that both PET tracers systematically underestimated tau pathology relative to neuropathology at preclinical and early symptomatic stages, with MK6240 demonstrating closer alignment than Flortaucipir. At the preclinical stage, Flortaucipir lagged neuropathology by approximately three Braak stages, whereas MK6240 lagged by approximately two stages. As cognitive impairment worsened, this offset progressively narrowed. At more severe cognitive decline stages, regional positivity frequencies converged across PET and neuropathology for both tracers, consistent with widespread neocortical tau involvement. The systematic underestimation at early stages is consistent with autopsy studies showing that Flortaucipir SUVRs in Braak I–III regions often fail to exceed young control levels even when histopathological tau is present(*15–17*). In contrast, the convergence between PET and neuropathology at more severe disease stages supports that tracer sensitivity constraints are most limiting during early tau accumulation but diminish as pathology becomes widespread. Together, these observations indicate that the relationship between PET-based and neuropathological Braak staging evolves across the course of disease and differs systematically by tracer, with tau PET measures substantially underestimating tau pathology burden during the earliest stages of pathology.

These findings have important clinical implications for the interpretation and application of tau PET imaging in AD(*41*). The substantial inter-tracer discordance at early and intermediate disease stages, precisely when therapeutic interventions are most likely to be effective(*42*), suggests that clinicians and researchers must exercise caution when comparing tau burden across studies using different tracers or when monitoring disease progression in individuals who switch between imaging agents. Furthermore, the systematic tendency of MK6240 to assign higher Braak stages than Flortaucipir may lead to different patient stratification decisions in clinical trials and different treatment recommendations in clinical practice. In addition, the observation that both tracers substantially lag behind neuropathological Braak stage frequencies at mild clinical severity suggests that individuals classified in vivo as Braak I–II may already harbor more advanced neocortical tangle pathology at autopsy. Clinically, this implies that tau PET staging may underestimate true pathological burden during the earliest symptomatic phases, potentially delaying recognition of biologically advanced disease, influencing prognostic discussions, and affecting optimal timing of therapeutic intervention. Taken together, these results underscore the need for tracer-specific normative data and staging criteria to ensure accurate patient classification and appropriate clinical interpretation.

A key strength of this study is that it presents the largest head-to-head comparison of tau PET tracers conducted to date, enabling this type of cross-tracer staging analysis for the first time. Nonetheless, many limitations should be acknowledged in our study. Tau PET provides an indirect measure of neurofibrillary pathology, and definitive validation of tracer-dependent Braak staging differences will require matched in vivo tau PET and postmortem histopathological assessment within the same individuals, particularly at early disease stages. Our cohort consists of self-selected individuals and was predominantly composed of White/Caucasian participants (94%), which may not be fully representative of the broader aging and AD populations and may limit the generalizability of these findings to more diverse populations. In addition, longitudinal studies with multiple time points are needed to determine how tracer-specific stage differences relate to disease progression, clinical decline, and therapeutic response over time.

In summary, this large head-to-head study demonstrates that tau PET-based Braak staging is hierarchically preserved across tracers but strongly tracer-dependent at the individual level. Systematic, directional differences between Flortaucipir and MK6240 are most pronounced at early and intermediate stages of AD and seem to reflect meaningful differences in biological sensitivity rather than methodological artifacts. These findings provide a framework for interpreting tau PET Braak stages across tracers and underscore the need for tracer-aware harmonization strategies in future research studies and clinical trials.

## Materials and Methods

### HEAD Study - Design and participants

The cross-sectional data analyzed in this study were derived from the HEAD Study (ClinicalTrials.gov identifier: NCT05361382) and included 644 older participants (50–90 years) and 38 young adults (18–28 years). HEAD is an ongoing multicenter, prospective, longitudinal study designed to enable head-to-head comparisons of tau PET tracers across the Alzheimer’s disease (AD) clinical continuum, encompassing cognitively unimpaired (CU) individuals, individuals with mild cognitive impairment (MCI), and individuals with dementia. The study is conducted across eight sites in the United States and Canada (Lawrence Berkeley National Laboratory, CA; University of California San Francisco, CA; Mayo Clinic, MN; Washington University in St. Louis, MO; University of Pittsburgh, PA; Brown University, RI; Houston Methodist Research Institute, TX; and McGill University, Canada). All participants undergo standardized clinical assessments, structural MRI, amyloid-β (Aβ) PET imaging, and head-to-head tau PET imaging with both [^18^F]Flortaucipir and [^18^F]MK6240. Additional information on the HEAD Study is available at https://head-study.info/home.

Eligibility criteria required the presence of a study partner capable of providing collateral information regarding functional status, the ability and willingness to undergo MRI and PET imaging, and fluency in an approved study language (English, Spanish, or French). Participants were excluded if they were unable to provide informed consent (or assent with proxy consent) or had medical conditions or medication use judged by site investigators to pose safety concerns or interfere with study procedures. Written informed consent was obtained from all participants or their legally authorized representatives, as applicable. Participants did not discontinue any medications to participate in the study. The study protocol was approved by the institutional review board at each participating site, with Advarra serving as the institutional review board of record for all U.S. centers (Protocol ID: Pro00061365). Participant sex was determined based on self-report and/or clinical records and was considered in the study design through inclusion of both male and female participants across the AD clinical continuum.

Clinical characterization was performed at enrolment using standardized instruments from the National Alzheimer’s Coordinating Center (NACC) Uniform Data Set (UDS; versions 3.0 or 4.0)(*43, 44*). These assessments included the Clinical Dementia Rating (CDR) scale and a comprehensive neuropsychological battery. Global cognitive status was evaluated using both the Montreal Cognitive Assessment (MoCA) and the Mini-Mental State Examination (MMSE). Participants were classified as cognitively unimpaired (CU, CDR Global = 0), mild cognitive impairment (MCI, CDR Global = 0.5), or dementia (CDR Global ≥1).

### HEAD Study - MRI acquisition and processing

Structural MRI data were acquired on 3T scanners (see **Supplementary Table 10** for scanners list) using harmonized acquisition protocols aligned with ADNI4 recommendations. High-resolution T1-weighted images were obtained in the sagittal plane using accelerated 3D MPRAGE or IRSPGR sequences.

Image preprocessing included correction for intensity inhomogeneity using N4 bias field correction, followed by tissue classification into grey matter, white matter, cerebrospinal fluid, skull, soft tissue, and air using the DARTEL framework implemented in SPM12. Whole-brain masks were generated by combining grey matter, white matter, and cerebrospinal fluid probability maps and applying a high- probability cut-off (>90%), with subsequent filling of internal cavities. Cerebellar segmentation was performed using FreeSurfer (v7.4.1) and FastSurfer (v2.1.1), incorporating regions defined by the SUIT atlas. Inferior cerebellar grey matter reference regions were delineated in native space and included bilateral Crus I/II, VIIb, VIIIa/b, IX, X, Vermis, and Vermis VII/VIII/IX. An additional whole-cerebellum reference region encompassing all SUIT-defined cerebellar areas was also generated. Spatial normalization of T1-weighted images to the ICBM 2020 Nonlinear Symmetric template was performed using ANTs with affine and symmetric diffeomorphic (SyN) transformations. All MRI-derived segmentations, reference regions, and spatial transformations were visually inspected for accuracy.

### HEAD Study - PET acquisition and processing

All participants underwent PET imaging with one Aβ tracer (where selection of tracer was dependent on site) and two tau PET tracers (see **Supplementary Table 10** for scanners and tracer list for each site). Radiotracers were administered as intravenous bolus injections following site-specific dosing guidelines. Aβ PET imaging was performed using [¹¹C]PiB (∼15 mCi), [¹□F]Florbetaben (∼8 mCi), or [¹□F]NAV4694 (∼8.1 mCi). Tau PET imaging consisted of paired acquisitions with [¹□F]MK6240 (∼5 mCi) and [¹□F]Flortaucipir (Tauvid; ∼10 mCi), performed on the same scanner within a 45-day interval.

PET data were reconstructed into four consecutive 5-minute frames using scanner-specific ordered-subset expectation maximization algorithms with standardized reconstruction settings and no post-reconstruction smoothing. Tracer-specific acquisition windows were as follows: [¹¹C]PiB (40–60 or 50–70 min), [^18^F]Florbetaben (90–110 min), [^18^F]NAV4694 (50–70 min), [^18^F]Flortaucipir (80–100 min), and [^18^F]MK6240 (90–110 min). Standard corrections for attenuation, motion, decay, scatter, random coincidences, and dead time were applied.

PET images were co-registered to native-space T1-weighted MRI using linear transformations implemented using Advanced Normalization Tools. Tau PET standardized uptake value ratios (SUVRs) were calculated using participant-specific inferior cerebellar grey matter reference regions, whereas Aβ PET SUVRs were calculated using a whole-cerebellum reference region in accordance with Centiloid calibration guidelines(*45*). MRI-derived spatial transformations were subsequently applied to bring PET images into stereotaxic space, and individual whole-brain masks were used to remove non-brain voxels. Final PET images were smoothed using scanner-specific Gaussian kernels to achieve an effective spatial resolution of 8 mm full width at half maximum. Anatomical regions of interest (ROIs) were defined using the CerebrA atlas, supplemented by a transentorhinal cortex segmentation generated using a validated protocol in 1-mm isotropic stereotaxic space.

Aβ burden was quantified in Centiloid units using a standard composite cortical ROI encompassing frontal, temporal, and parietal cortices, the precuneus, anterior striatum, and insula. Aβ positivity was defined as a Centiloid value ≥10.

Tau PET analyses were performed separately for each tracer using Braak-like ROIs defined according to Pascoal *et al.* ^4^ with optimization of the Braak II ROI to reduce off-target spillover: Braak I (transentorhinal cortex), Braak II (entorhinal cortex), Braak III (amygdala and parahippocampal, fusiform, and lingual gyri), Braak IV (inferior and middle temporal cortices, anterior and posterior cingulate, isthmus cingulate, and insula), Braak V (lateral occipital, inferior and superior parietal cortices, superior and transverse temporal gyri, precuneus, supramarginal gyrus, and inferior frontal regions), and Braak VI (precentral, postcentral, paracentral, and pericalcarine cortices).

Regional tau tracer uptake for each ROI was calculated as the median SUVR across all voxels within that region. Tau positivity cut-offs were defined for each tracer and ROI as the mean SUVR + 2.0 standard deviations (S.D.) derived from a young adult reference group accordingly with Pascoal *et al.*(*11*). Young participants were used exclusively for cut-off derivation and were excluded from all subsequent analyses. To assess the robustness of our findings to threshold selection, we additionally evaluated alternative cut-off strategies, including: reference-based methods using either a cognitively unimpaired amyloid-negative (CU Aβ-) group (aged ≤65 years, Centiloid < 10) or young adults (aged 18–28 years) at varying S.D. thresholds (mean + 1.5, 1.8, 2.0, or 2.5 S.D.); data-driven Gaussian mixture modeling (GMM); 95th percentile of the CU Aβ- group (aged ≤65 years, Centiloid < 10) reference distribution; hybrid combinations applying different threshold methods to Flortaucipir versus MK6240; and selected methods with non-optimized Braak II ROI.

Participants were assigned the highest Braak stage for which all preceding ROIs met tracer- specific positivity criteria. Individuals exhibiting positivity in later-stage ROIs in the absence of positivity in one or more earlier ROIs were classified as Braak stage “non-conformant” whereas individuals following the hierarchical progression of Braak were classified as “conformant”. For the agreement by Braak stage analyses, agreement between Flortaucipir and MK6240 stage assignments was evaluated using a stage-specific overlap metric analogous to a Venn diagram intersection-over-union approach. For each Braak stage, the numerator corresponded to the number of individuals assigned to the same stage by both tracers, whereas the denominator corresponded to the total number of individuals assigned to that stage by either tracer.

To characterize voxel-level tau abnormalities, abnormal voxel maps were generated separately for Flortaucipir and MK6240 by binarizing individual PET images using voxel-wise cut-offs defined as the mean SUVR + 2.0 S.D. from the young reference group, calculated independently for each tracer. These binarized maps were subsequently overlaid to identify voxels classified as abnormal by Flortaucipir only, MK6240 only, or by both tracers within the same individual. Frequency-of-abnormal-voxel maps were generated for each tracer by averaging the binarized abnormal voxel maps across individuals within each group and tracer, yielding voxel-wise representations of the proportion of participants exhibiting abnormal tau signal in that group. Z-score maps were also generated at the voxel level for each tracer. For each voxel, Z-scores were computed by subtracting the mean SUVR of the young reference group from the individual SUVR value and dividing by the corresponding S.D. of the young reference group.

Probability-based harmonization matrices for tau-PET Braak stage conversion were generated by tabulating all observed MK6240 and Flortaucipir Braak stage combinations. For each conversion direction, probabilities were calculated by dividing the number of individuals assigned to a given combination of MK6240 and Flortaucipir stages by the total number of individuals assigned to the predictor stage. These probabilities were used to generate stage-conversion tables under both the 7-class and 4-class Braak staging frameworks. Non-conformant individuals were excluded from these analyses.

Probability-based conversion matrices mapping tau-PET to neuropathological Braak stages were generated separately for MK6240 and Flortaucipir using Braak stage distributions from the HEAD and NACC cohorts. Within each CDR-SB severity bin, the frequency distributions of tau PET and neuropathological Braak stages were computed independently. For each tau PET Braak stage, the corresponding neuropathological stage was identified by matching cumulative frequency ranks across the two distributions. Conditional probabilities of assignment to each neuropathological stage were then derived using Gaussian kernels centered on the matched stage, weighted by the observed neuropathological stage frequencies, and normalized to sum to 100% within each tau PET stage. Conversion matrices were generated under both the 7-class and 4-class Braak staging frameworks. Non- conformant individuals were excluded from these analyses.

### HEAD Study - Blood collection and measurements

Blood samples were drawn into commercial ethylenediaminetetraacetic acid (EDTA) tubes in accordance with NCRAD guidelines. Plasma samples were aliquoted, stored at -80 °C, and transferred under strict temperature-controlled conditions to the University of Pittsburgh, where they were thawed for the first time. Plasma p-tau217 was measured using the ALZpath Advantage PLUS assay (#104570), and GFAP was measured using the Neurology 4-Plex E Advantage PLUS assay (#104465), both on the single- molecule array (Simoa) platform (Quanterix®).

### NACC Cohort - Participants and Data

Clinical and neuropathologic data were drawn from the NACC cohort, a centralized repository that curates longitudinal data contributed by Alzheimer’s Disease Research Centers (ADRCs) supported by the U.S. National Institute on Aging. All contributing ADRCs were required to obtain written informed consent from their participants and obtain approval from their institutional review board before submitting data to the NACC. These centers prospectively follow participants using harmonized assessment protocols, with clinical, cognitive, and demographic information compiled within the Uniform Data Set (UDS)(*43, 44*). Participants who elected brain donation underwent postmortem examination according to standardized neuropathological procedures, with findings recorded in the NACC Neuropathology Data Set (NPDS). The present study included 4,422 individuals from the NACC autopsy cohort (mean age, 80.0 ± 11.5 years) who had both neuropathologic data and at least one CDR assessment obtained within one year prior to death. Detailed descriptions of NACC data acquisition, harmonization, neuropathological assessments and quality assurance procedures are available elsewhere(*46, 47*). Participants were classified as CU (CDR Global = 0), MCI (CDR Global = 0.5), or dementia (CDR Global ≥1). Neuropathological Aβ positivity was defined as a CERAD value ≥ 2, shown previously to be equivalent to Centiloid ≥ 10 in Aβ PET ^7^

### Statistical analysis

For demographic, clinical, genetic, and biomarker characteristics (**Table 1**, **Supplementary Table 1 and Supplementary Table 2**), categorical variables were compared across clinical groups using Pearson’s χ² tests or Fisher’s exact tests, as appropriate, and continuous variables using Kruskal-Wallis tests. When overall effects were significant, post hoc pairwise comparisons were performed using Fisher’s exact tests (categorical variables) or Wilcoxon rank-sum tests (continuous variables), with false discovery rate (FDR) correction applied. Braak stage conformity and non-conformity rates were compared using χ² tests. Differences in Braak stage frequency between Flortaucipir and MK6240 (**Supplementary Table 3**) were assessed using stage-specific χ² tests or Fisher’s exact tests within clinical and Aβ-status strata. One test was performed per Braak stage, and FDR correction was applied across stage-wise comparisons. Inter- tracer Braak stage agreement was quantified using Cohen’s κ statistics (unweighted and linearly weighted). Group-averaged and voxel-wise tau PET analyses (SUVR maps, frequency-of-abnormal-voxel maps, and Z-score maps) were descriptive and not subjected to voxel-wise inferential testing. To assess the biological and clinical relevance of tracer-dependent Braak staging differences, individuals assigned a higher Braak stage by MK6240 than by Flortaucipir (MK6240-upstaged) were compared with concordantly staged individuals within each Flortaucipir-defined Braak stage. For each stage, biomarker and cognitive measures were compared between groups using Welch two-sample t-tests. This analysis was restricted to the 4-class scheme due to the limited number of individuals in some groups in the 7- class scheme. Comparisons between neuropathological Braak stage and PET-based Braak staging were performed separately for each tau PET tracer and CDR-SB group. For each Braak ROI, the proportion of tau-positive individuals was compared between neuropathology and PET using χ² tests or Fisher’s exact tests. FDR correction was applied across Braak ROIs (**Supplementary Tables 8 and 9**). To assess the robustness of PET-to-neuropathology comparisons to potential demographic confounding, a sensitivity analysis was performed using 1:1 exact matching of PET and neuropathology participants on age, sex, and CDR-SB. Matched samples were used to repeat the proportion comparisons between tau PET and neuropathological Braak ROI positivity frequencies.

## Data Availability

All data produced in the present study are available upon reasonable request to the authors.

## Acknowledgements

We thank all HEAD Study personnel for their dedication and all participants, as well as their families and caregivers, for their time, data, biological specimens, and continued support. We are grateful to the investigators, coordinators, and research staff across participating sites for their essential contributions to participant recruitment, study coordination, and data collection, and to the referring clinicians and community partners who facilitated recruitment efforts.

The HEAD Study was funded by the National Institute on Aging (R01AG073267). This work was also supported by the Alzheimer’s Association (AARFD-24-1307665 to A.R.; AARFD-22-974627 to B.B.; AARFD-24-1243899 to G.P.; AARFD-23-1150249 to G.N.-B.; AARFD-22-923814 to P.C.L.F.; AARFD-24-1313939 to M.S.R.). Imaging and data repository infrastructure were supported by the Laboratory of Neuroimaging (LONI), which is funded by the National Institutes of Health through the National Institute of Biomedical Imaging and Bioengineering (P41EB015922). We also acknowledge the National Centralized Repository for Alzheimer’s Disease and Related Dementias (NCRAD) for biospecimen storage and APOE genotyping, supported by the National Institute on Aging under cooperative agreement U24AG021886. Finally, we thank Avid Radiopharmaceuticals and Lantheus/Cerveau Technologies for enabling use of the Flortaucipir and MK6240 radiotracers, respectively.

We acknowledge the National Alzheimer’s Coordinating Center (NACC) for access to the Uniform Data Set. The NACC database is funded by NIA/NIH Grant U24AG072122. NACC data are contributed by the NIA-funded ADRCs: P30 AG062429 (PI James Brewer, MD, PhD), P30 AG066468 (PI Oscar Lopez, MD), P30 AG062421 (PI Teresa Gomez-Isla, MD), P30 AG066509 (PI Thomas Grabowski, MD), P30 AG066514 (PI Mary Sano, PhD), P30 AG066530 (PI Helena Chui, MD, Arthur Toga, PhD), P30 AG066507 (PI Marilyn Albert, PhD), P30 AG066444 (PI David Holtzman, MD), P30 AG066518 (PIs Lisa Silbert, MD, Kevin Duff, PhD), P30 AG066512 (PI Thomas Wisniewski, MD), P30 AG066462 (PI Scott Small, MD), P30 AG072979 (PI David Wolk, MD), P30 AG072972 (PIs Charles DeCarli, MD, Rachel Whitmer, PhD), P30 AG072976 (PI Andrew Saykin, PsyD), P30 AG072975 (PI Julie Schneider, MD, MS), P30 AG072978 (PI Ann McKee, MD), P30 AG072977 (PI Robert Vassar, PhD), P30 AG066519 (PI Joshua Grill, PhD), P30 AG062677 (PIs Brad Boeve, MD, Ronald Petersen, MD, PhD), P30 AG079280 (PI Jessica Langbaum, PhD), P30 AG062422 (PI Gil Rabinovici, MD), P30 AG066511 (PI Allan Levey, MD, PhD), P30 AG072946 (PI Linda Van Eldik, PhD), P30 AG062715 (PI Sanjay Asthana, MD, FRCP), P30 AG072973 (PI Russell Swerdlow, MD), P30 AG066506 (PIs Glenn Smith, PhD, ABPP, David Lowenstein, PhD, Ranjan Duara, MD), P30 AG066508 (PIs Stephen Strittmatter, MD, PhD, Christopher Van Dyck, MD), P30 AG066515 (PI Victor Henderson, MD, MS), P30 AG072947 (PI Suzanne Craft, PhD), P30 AG072931 (PI Henry Paulson, MD, PhD), P30 AG066546 (PIs Sudha Seshadri, MD, Gladys Maestre, MD, PhD), P30 AG086401 (PI Erik Roberson, MD, PhD), P30 AG086404 (PI Gary Rosenberg, MD), P30 AG086403 (PI Angela Jefferson, PhD), P30 AG072958 (PIs Heather Whitson, MD, Gwenn Garden, MD, PhD), P30 AG072959 (PI Jagan Pillai, MD, PhD), P30 AG092752 (Ihab Hajjar, MD, MS).

## Data availability

Request for raw or analyzed data and materials for the HEAD study should be directed to the corresponding author. All requests will be reviewed by the investigators and participating institutions to determine whether they are subject to intellectual property or confidentiality restrictions. De-identified data will be made accessible to qualified academic researchers upon request, specifically for the purpose of replicating the methods and findings outlined in this paper. Any releasable data and materials will be provided under a material transfer agreement. Data from the NACC study are publicly available in the NACC repository https://www.naccdata.org/data-request-process/. The codes used for the data analyses in the present work will be made available from the corresponding author upon request.

## Author contributions

T.A.P., S.B., .R.-N., B.A.G., B.P., H.O., V.L., D.N.S.-M., and J.F. conceived and designed the study. A.R. developed the analytical methodology, conducted the analyses and drafted the manuscript. T.A.P., A.R., C.S., E.R., and M.S.M. performed the interpretation of the results. D.L.T. oversaw the statistical analyses. Data acquisition was carried out by T.A.P., S.B., P.R.-N, B.A.G., B.P., H.O., V.L., D.N.S.-M., J.F., J.M. and F.Z.L. Image processing and data quality control were performed by G.P., G.B.-N., L.A., F.Z.L., B.B., P.C.L.F., A.R., E.R., and M.S.M. Plasma biomarker measurements were performed by P.C.L.F, A.R. and B.B. All authors contributed to manuscript revision and approved the final version.

