## Supplementary Information for "Tau PET Braak staging across the Alzheimer Disease continuum using Flortaucipir and MK6240"

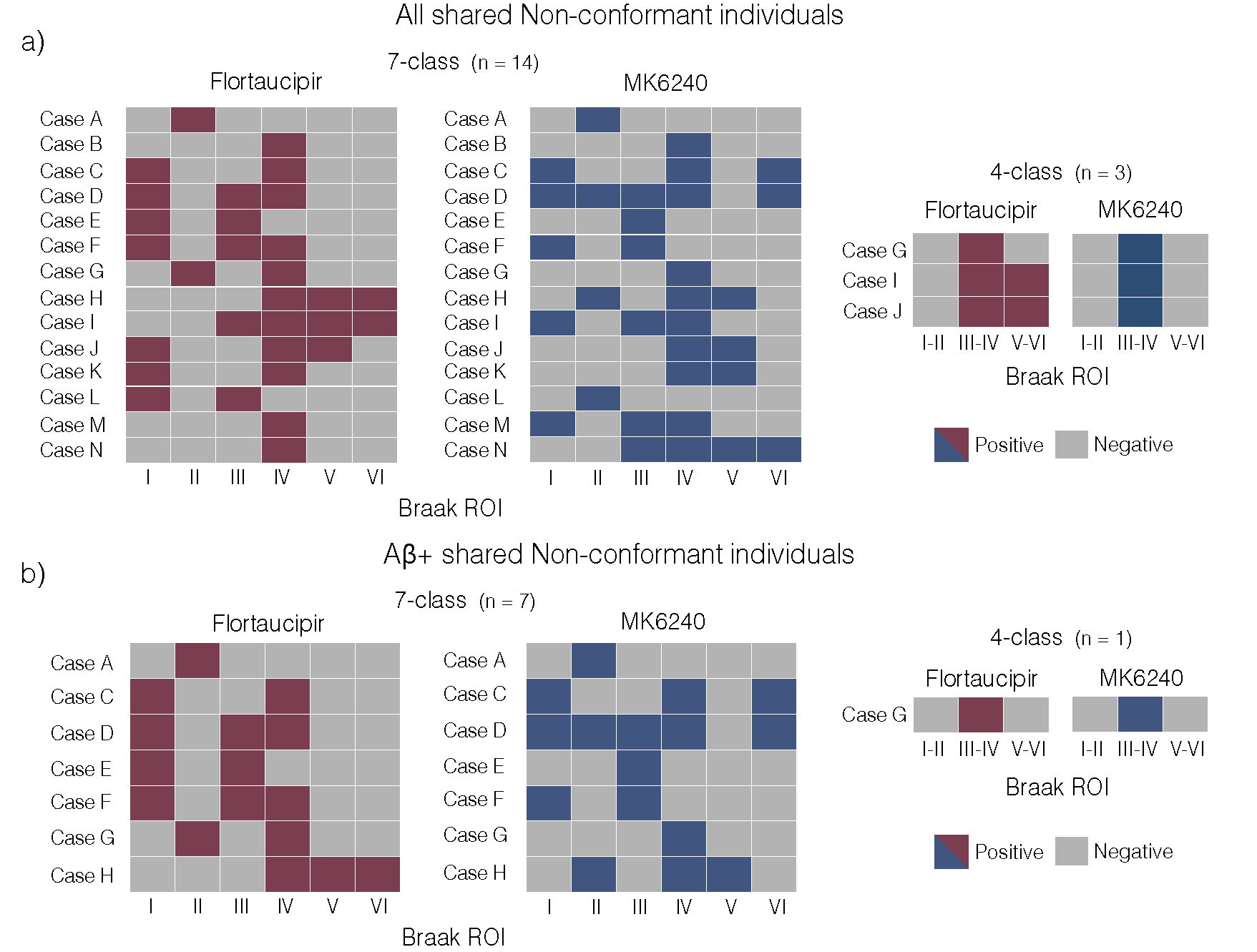

**Supplementary Figure 1. Flortaucipir and MK6240 shared Braak non-conformant cases.**

**(a,b)** Binary matrices showing Braak regional tau positivity patterns for individuals classified as Braak non-conformant by both Flortaucipir and MK6240 under the 7-class and 4-class Braak staging schemes. Each row represents an individual case, and each column represents a Braak region of interest (ROI). Results are shown for all shared non-conformant individuals **(a)** and for the subset of Aβ+ shared non-conformant individuals **(b).** Non-conformant cases represent individuals whose tau distribution did not follow the hierarchical Braak ordering.

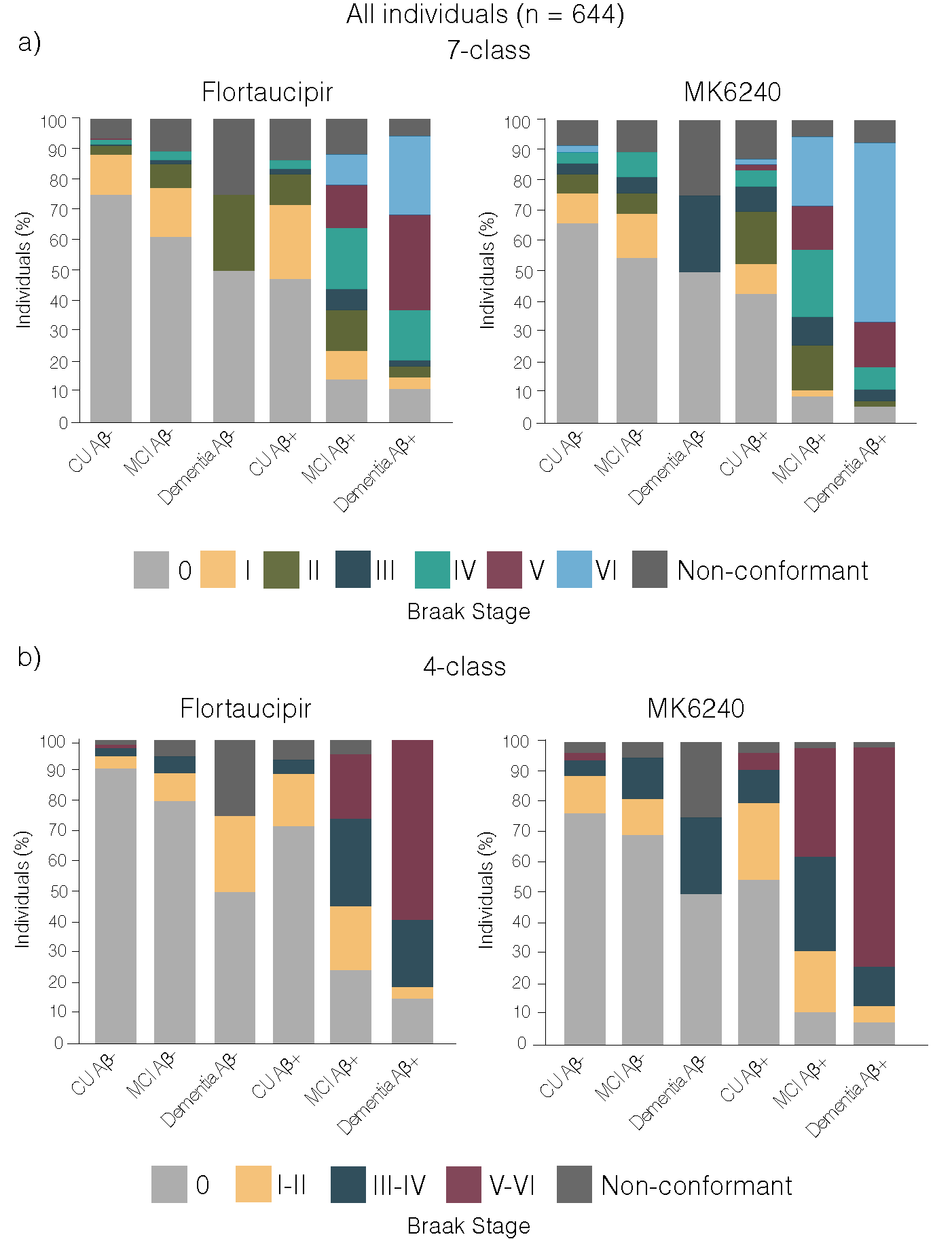

**Supplementary Figure 2. Braak stage distributions across clinical groups and Aβ status for Flortaucipir and MK6240.**

Distribution of Braak stages stratified by clinical group and Aβ status for each tracer under the 7-class (**a**) and 4-class (**b**) staging schemes. (N = 644). Non-conformant cases represent individuals whose tau distribution did not follow the hierarchical Braak ordering.

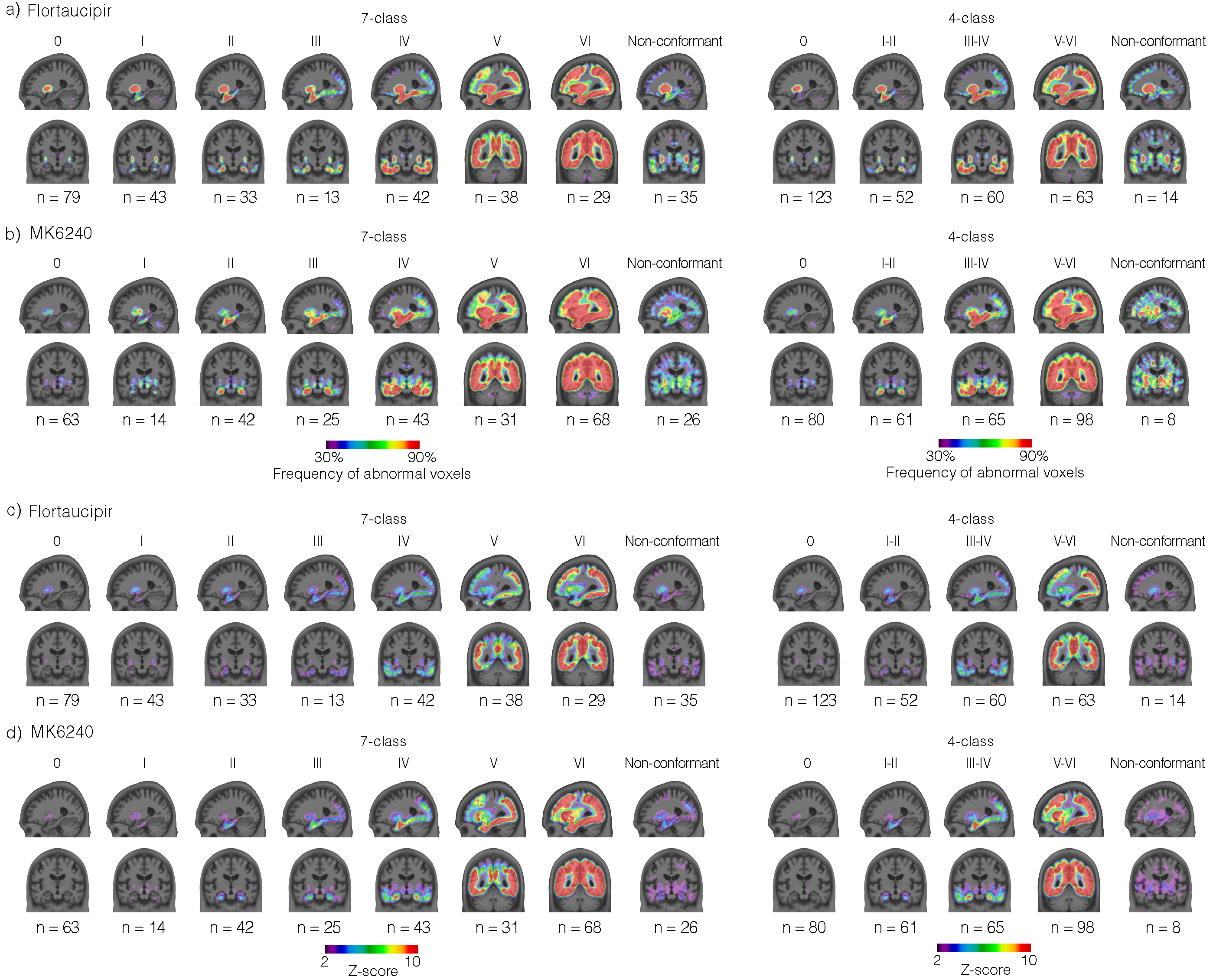

**Supplementary Figure 3. Voxel-wise tau PET patterns across Braak stages.**

Group-averaged brain maps showing the frequency of abnormal voxels (**a,b**) and Z-score (**c,d**) brain maps across Braak stages for Flortaucipir and MK6240**.** Abnormal voxels were defined using voxel-wise cut-offs corresponding to mean SUVR + 2.0 S.D. derived from the young reference group for each tracer at the voxel level. Z-scores were calculated as the deviation from the mean SUVR of the young reference group divided by the S.D. at each voxel. For all panels, representative sagittal and coronal slices are shown. All results are presented under the 7-class and 4-class Braak staging schemes. All analyses were restricted to Aβ+ individuals (Centiloid ≥ 10; N = 312). Non-conformant cases represent individuals whose tau distribution did not follow the hierarchical Braak ordering and are shown separately.

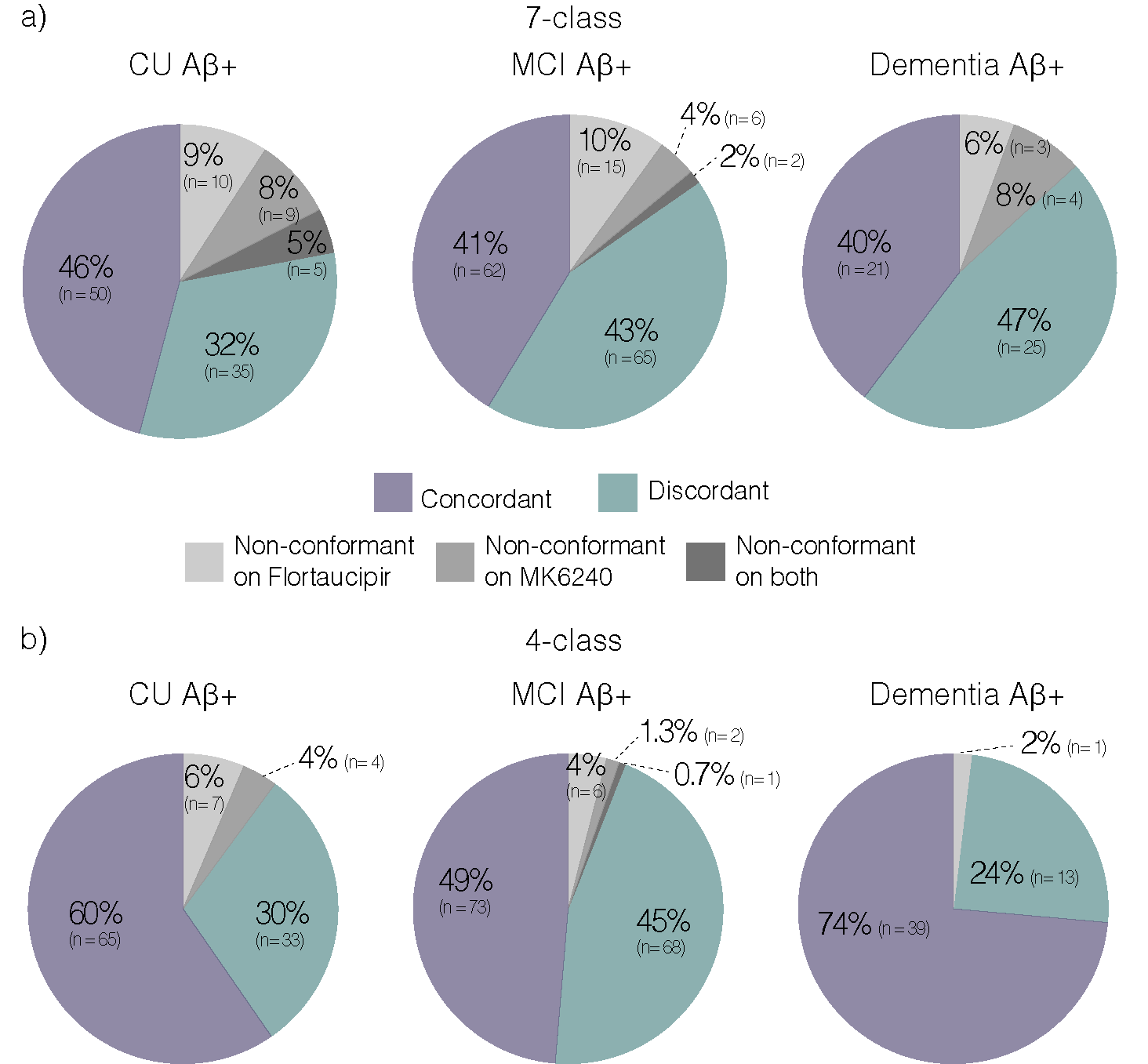

**Supplementary Figure 4. Inter-tracer agreement in Braak stage assignment by clinical status.
(a-b)** Proportion of individuals classified as Braak concordant, Braak discordant, or Braak non-conformant between Flortaucipir and MK6240, stratified by clinical status (CU, MCI, and dementia). Results are shown under the 7-class (**a**) and 4-class (**b**) Braak staging schemes. Concordant = identical Braak stage assigned by both tracers; discordant = different Braak stage assigned; non-conformant = hierarchical Braak ordering not followed by one or both tracers. All analyses were restricted to Aβ+ individuals (Centiloid ≥ 10; N = 312).

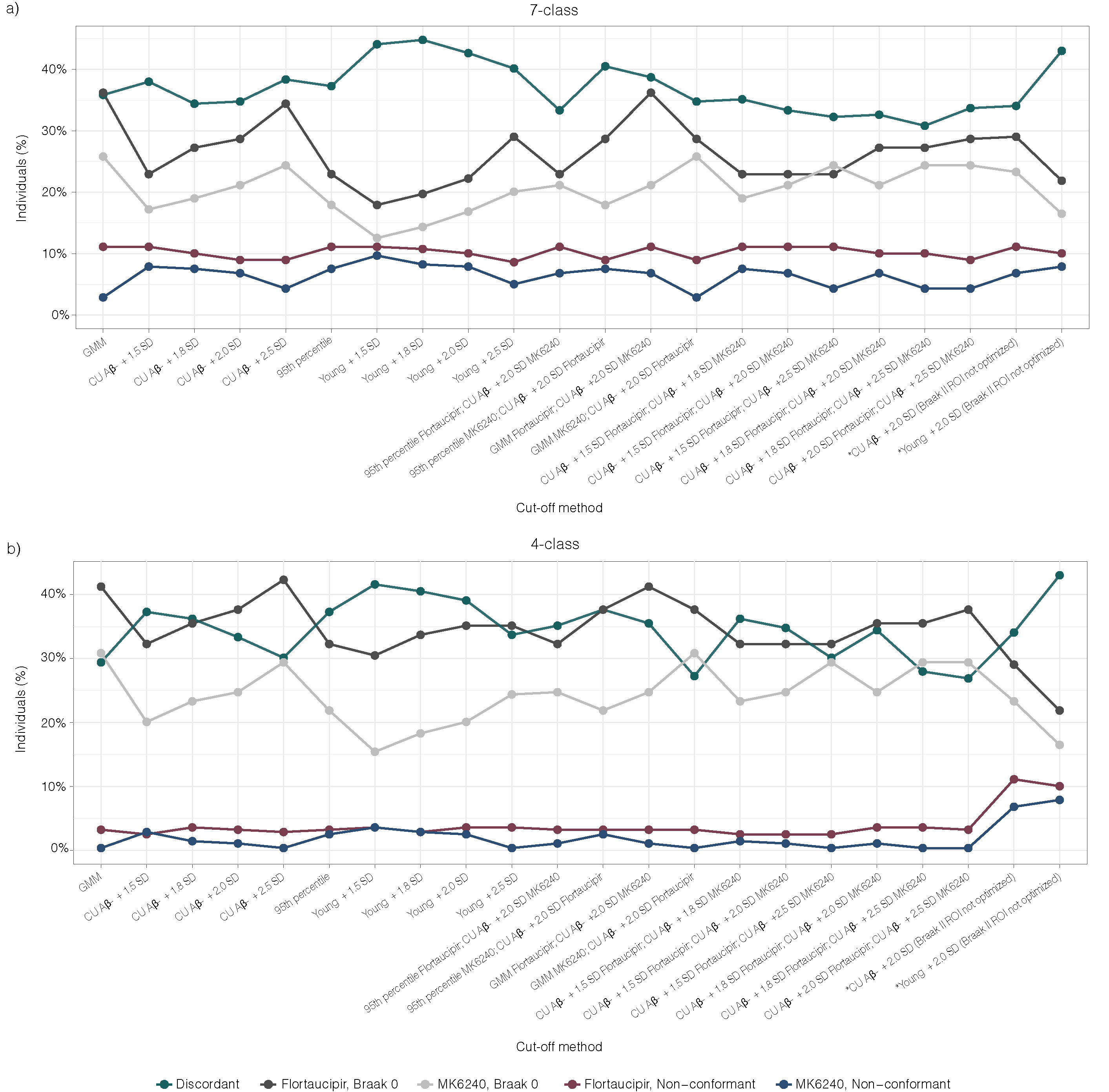

**Supplementary Figure 5. Braak stage classification across different positivity cut-offs.**

**(a,b)** Proportion of individuals classified as Braak discordant, Braak non-conformant, or Braak 0 across alternative tau PET positivity cut-offs and cut-off combinations for Flortaucipir and MK6240. Results are shown under the 7-class **(a)** and 4-class **(b)** Braak staging schemes. All analyses were restricted to Aβ+ individuals (Centiloid ≥ 10; N = 312). Cut-off methods include: CU Aβ- reference group (aged ≤65 years) SUVR mean + 1.5, 2.0, or 2.5 S.D.; young control group mean SUVR + 1.5, 2.0, or 2.5 S.D.; Gaussian mixture modeling (GMM); 95th percentile of CU Aβ- reference distribution; and combinations of these cut-offs applied differentially to Flortaucipir versus MK6240. Methods marked with asterisks indicate non-optimized Braak II ROI. Non-conformant cases represent individuals whose tau distribution did not follow the hierarchical Braak ordering.

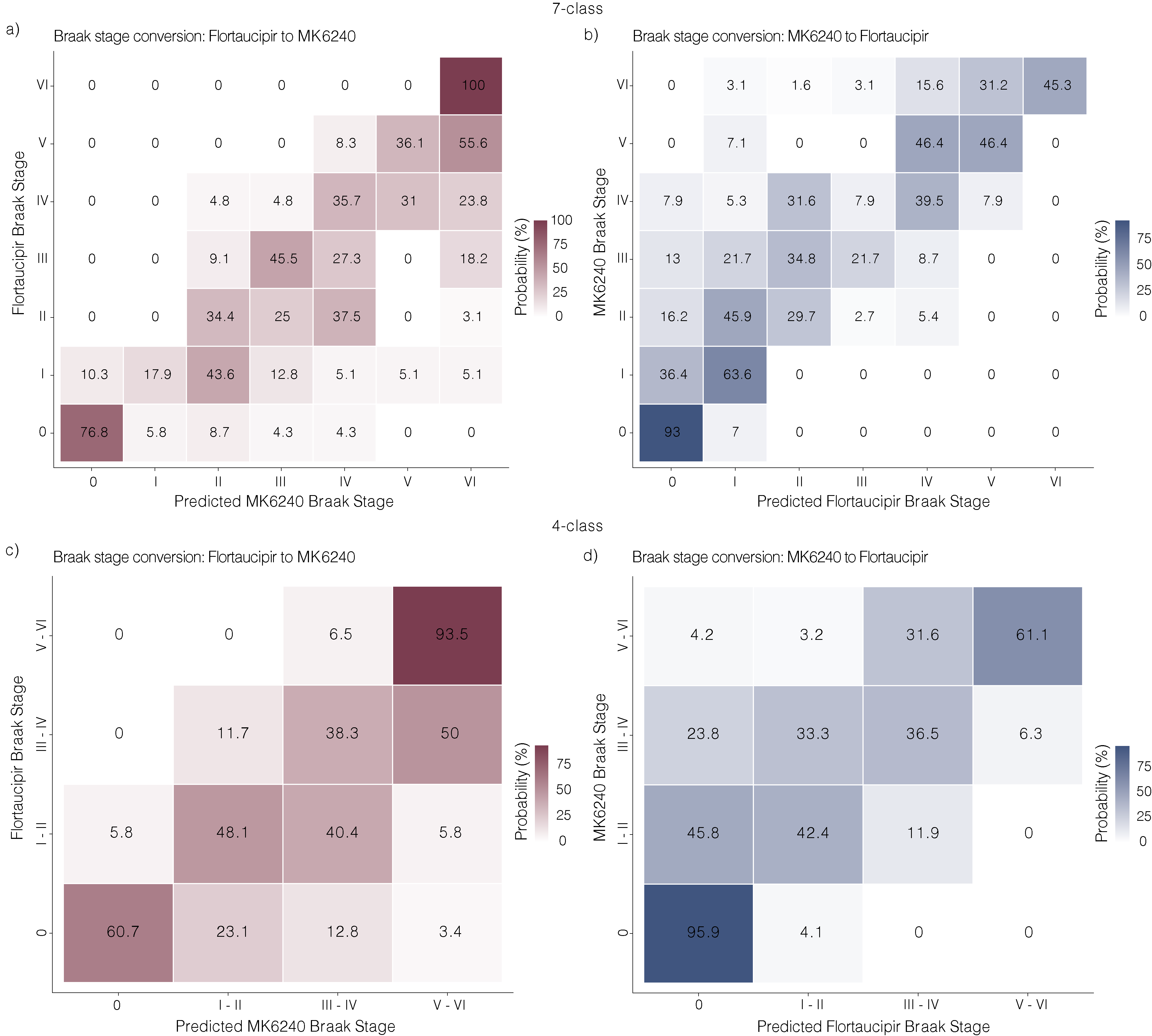

**Supplementary Figure 6. Cross-tracer Braak stage conversion probabilities.**

Probability-based harmonization matrices for Braak stage conversion between Flortaucipir and MK6240 under the 7-class (**a, b**) and 4-class (**c, d**) staging schemes. Panels (**a**) and (**c**) show the probability of each corresponding MK6240 Braak stage given a Flortaucipir-defined stage, and panels (**b**) and (**d**) show the reverse. Each cell represents the probability (%) that a given stage in one tracer corresponds to a given stage in the other, with rows summing to 100%. All analyses were restricted to Aβ+ individuals (Centiloid ≥ 10) and excluded Braak non-conformant cases.

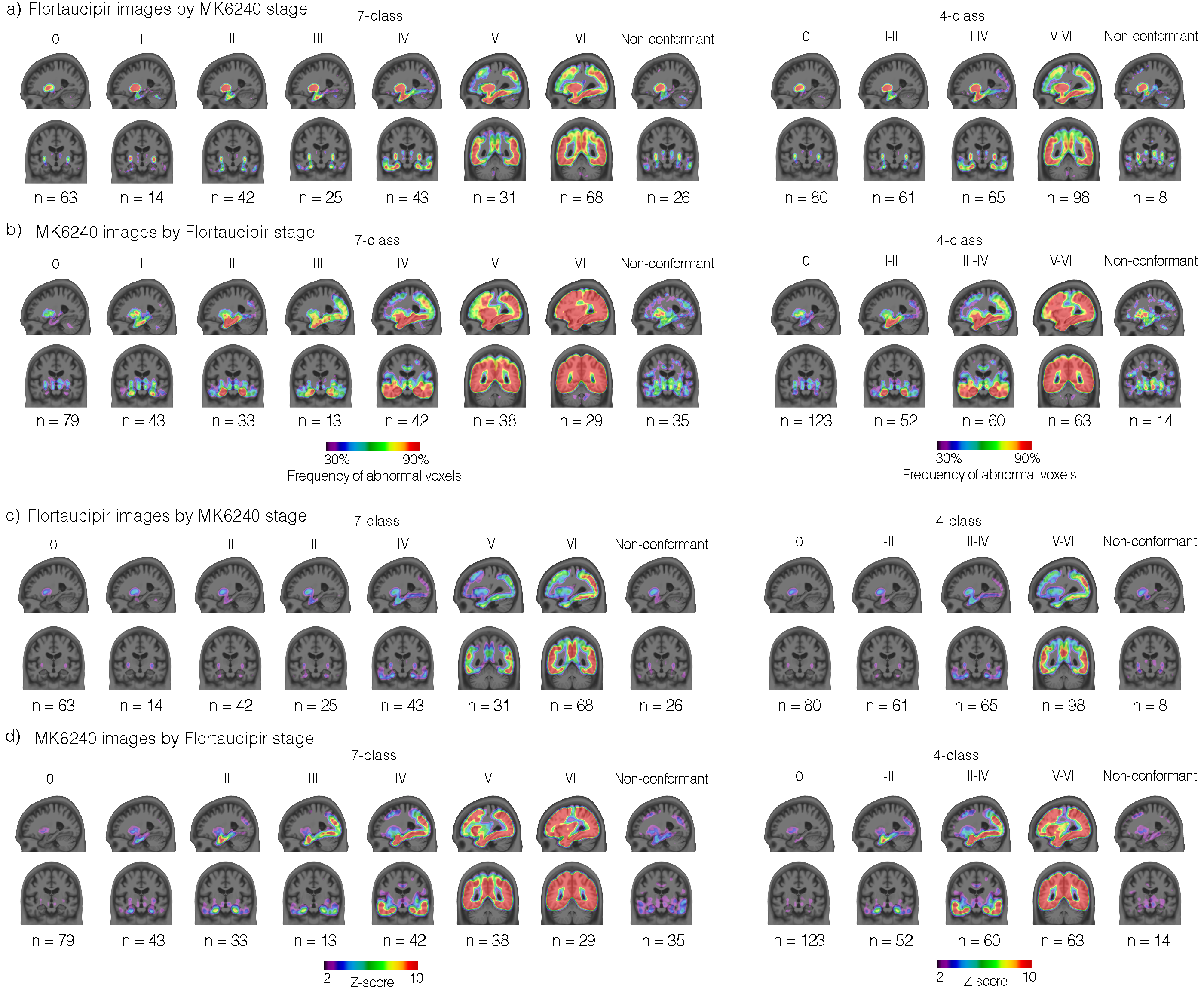

**Supplementary Figure 7. Voxel-wise tau PET patterns stratified by cross-tracer Braak stage assignment.**

**(a)** Group-averaged frequency-of-abnormal-voxel maps for Flortaucipir stratified by Braak stage as assigned by MK6240. **(b)** Group-averaged frequency-of-abnormal-voxel maps for MK6240 stratified by Braak stage as assigned by Flortaucipir. **(c)** Group-averaged Z-score maps for Flortaucipir stratified by MK6240-assigned Braak stage. **(d)** Group-averaged Z-score maps for MK6240 stratified by Flortaucipir-assigned Braak stage. Abnormal voxels were defined using voxel-wise cut-offs corresponding to mean SUVR + 2.0 S.D. derived from the young reference group for each tracer. Z-scores were calculated as the deviation from the mean SUVR of the young reference group divided by the S.D. at each voxel. For all panels, representative sagittal and coronal slices are shown. All results are presented under the 7-class and 4-class Braak staging schemes. All analyses were restricted to Aβ+ individuals (Centiloid ≥ 10; N = 312). Non-conformant cases represent individuals whose tau distribution did not follow the hierarchical Braak ordering and are shown separately.

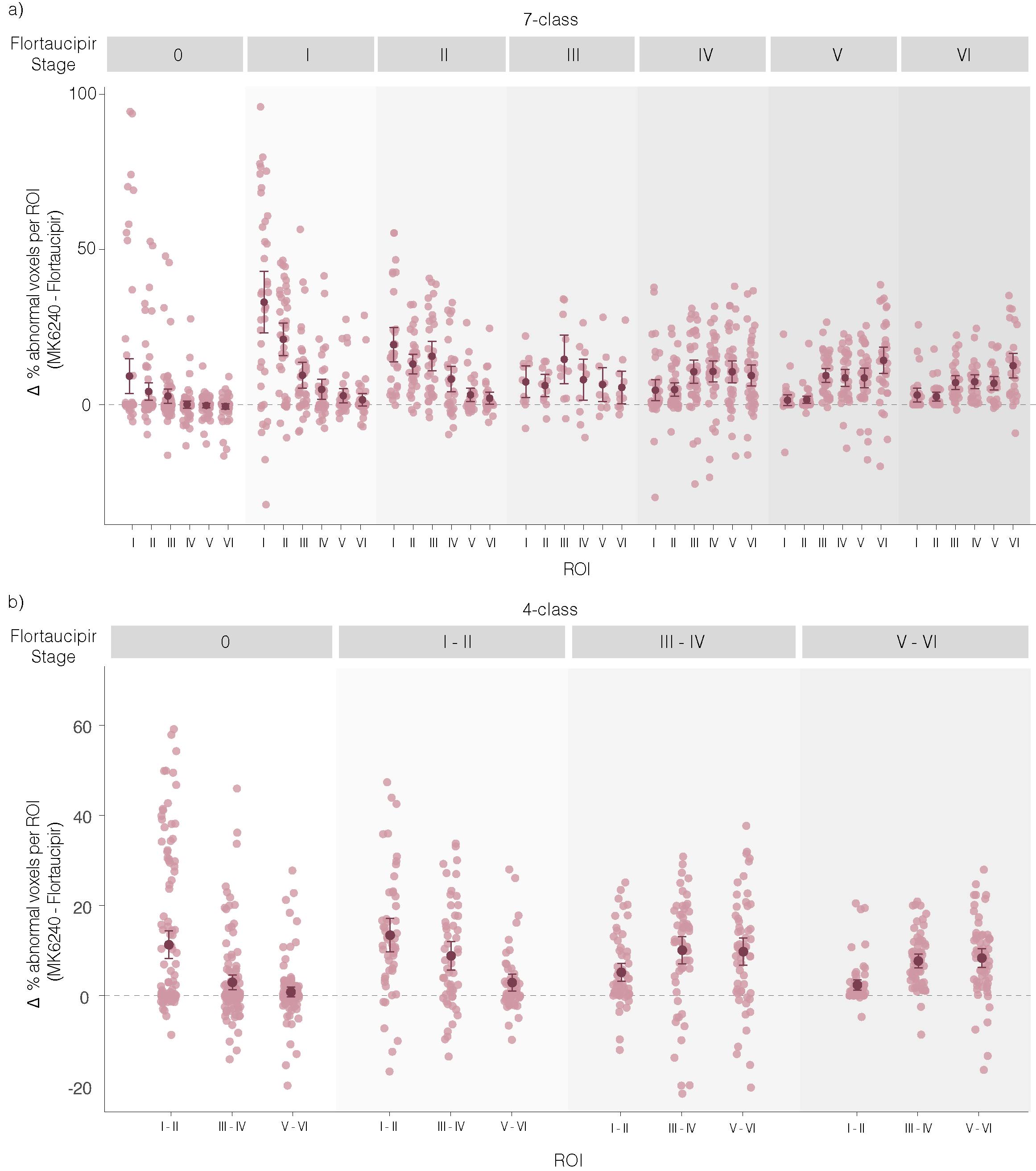

**Supplementary Figure 8. Regional differences in abnormal voxel coverage between MK6240 and Flortaucipir.**

Differences in the proportion of abnormal voxels within Braak ROIs between MK6240 and Flortaucipir (Δ = MK6240 − Flortaucipir) across Flortaucipir-defined Braak stages in Aβ+ individuals (N = 312) under the 7-class (**a**) and 4-class (**b**) staging schemes. Each pink point represents an individual participant, while burgundy points and error bars indicate the mean and 95% confidence interval within each ROI and stage. Positive values indicate a greater proportion of abnormal voxels detected by MK6240 relative to Flortaucipir.

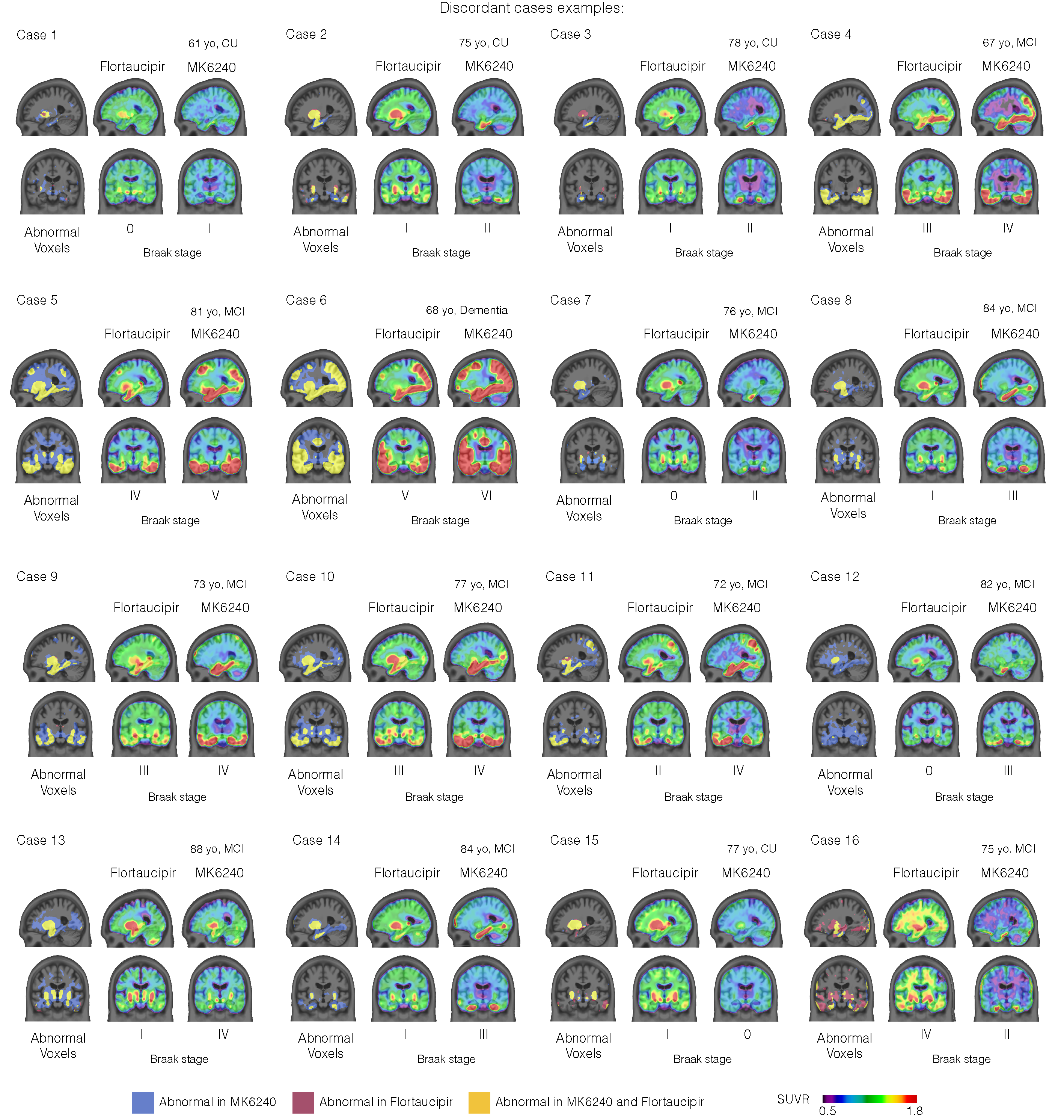

**Supplementary Figure 9. Representative discordant case examples.**

Representative cases illustrating discordant Braak stage assignments between Flortaucipir and MK6240. For each case, tau PET SUVR and corresponding abnormal voxel brain maps are shown for both tracers. Abnormal voxels were defined using voxel-wise cut-offs corresponding to mean SUVR + 2.0 S.D. derived from the young reference group for each tracer at the voxel level. Regions classified as abnormal by MK6240 only, Flortaucipir only, or by both tracers are indicated. Representative sagittal and coronal slices are shown. All cases represented are Aβ+ individuals (Centiloid ≥ 10).

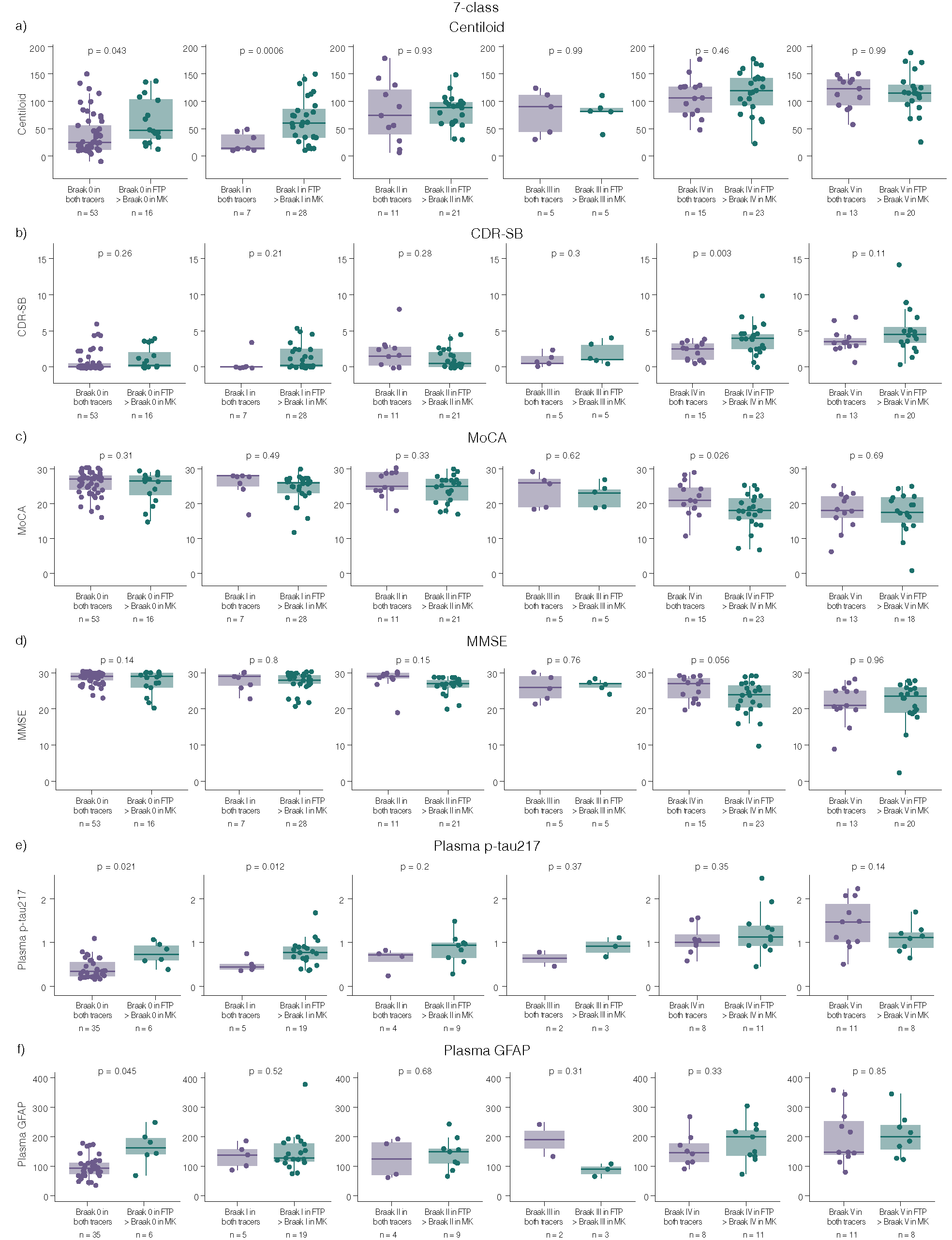

**Supplementary Figure 10. Higher MK6240-based Braak staging identifies individuals with greater biological and cognitive severity.**

Distributions of Aβ burden (Centiloid; **a**), cognitive performance (CDR-SB, MoCA and MMSE; **b**, **c, d**), and plasma biomarkers (p-tau217 and GFAP; **e**, **f**) in Aβ+ individuals grouped by Flortaucipir-based Braak stage and further stratified by whether MK6240 assigned the same stage or a stage higher classification under the 7-class Braak staging scheme. All analyses were restricted to Aβ+ individuals (Centiloid ≥ 10). Points represent individual participants. Boxes indicate the interquartile range (25th–75th percentile), the central line represents the median, and whiskers extend to 1.5 × the interquartile range. Individuals classified as Braak non-conformant and those assigned to a higher Braak stage by Flortaucipir than MK6240 were excluded from this analysis (N = 277). FTP = Flortaucipir; MK = MK6240. CDR-SB = Clinical Dementia Rating Sum of Boxes; MMSE = Mini-Mental State Examination; MoCA = Montreal Cognitive Assessment.

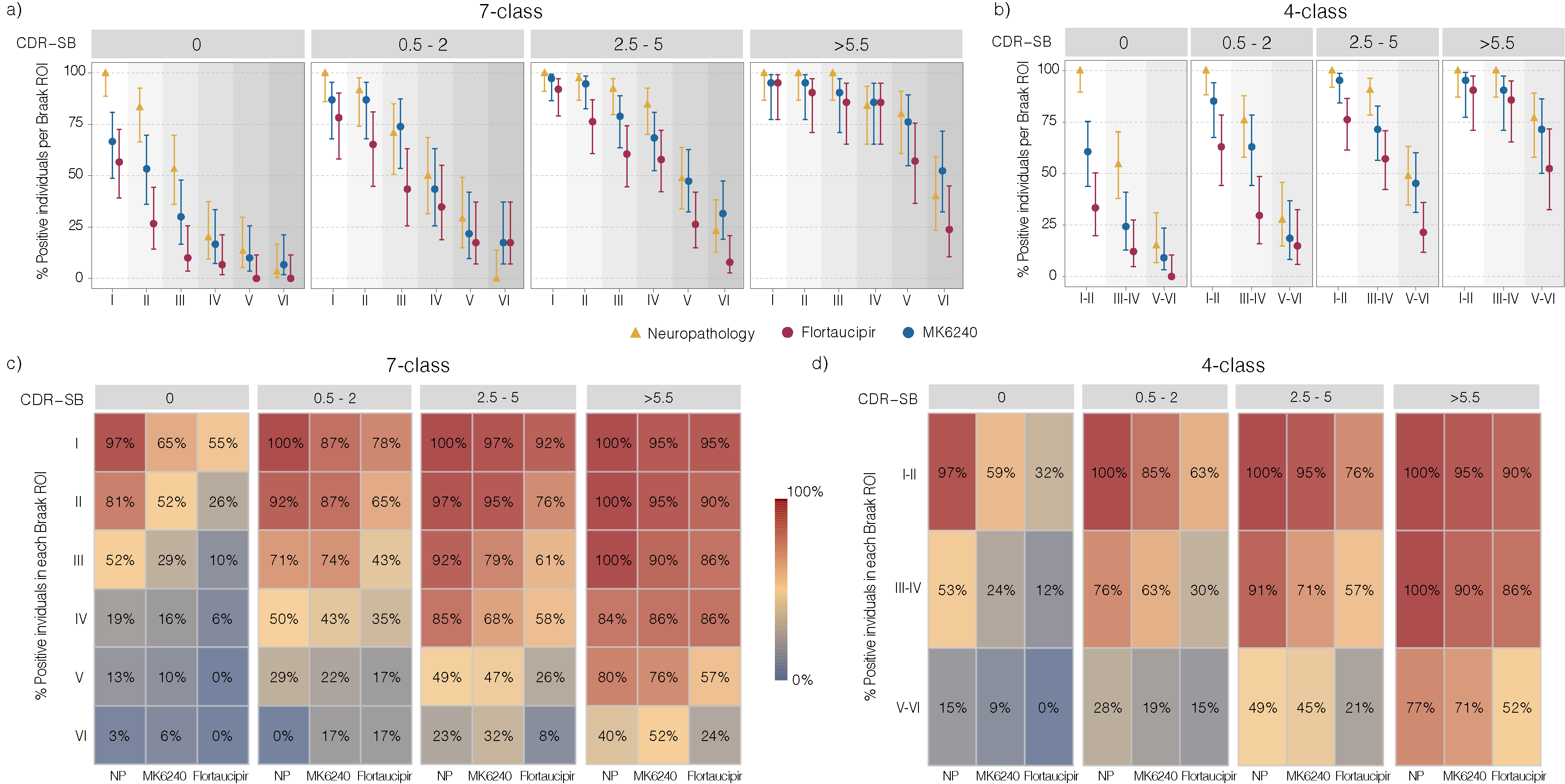

**Supplementary Figure 11. Mismatch between tau PET and neuropathological Braak stage frequencies across the AD spectrum in a matched samples analysis.**

Dot plots and heat maps representing the proportion of tau-positive individuals in each Braak ROI stratified by cognitive performance using CDR-SB categories under the 7-class (**a, c**) and 4-class (**b, d**) Braak staging schemes. Neuropathology data are derived from the NACC cohort and PET data from the HEAD Study cohort. Analyses were performed in 1:1 matched samples based on age, sex, and CDR-SB to ensure comparable groups across datasets. Dots represent group-level proportions and error bars the 95% confidence interval. All analyses were restricted to Aβ+ individuals (CERAD 2/3 for neuropathology or Centiloid ≥ 10 for PET) and excluded Braak non-conformant cases. Sample sizes per CDR-SB category for both neuropathology and PET: 7-class 0 (N = 30), 0.5–2 (N = 23), 2.5–5 (N = 38), >5.5 (N = 21); 4-class 0 (N = 33), 0.5–2 (N = 27), 2.5–5 (N = 42), >5.5 (N = 21). NP = neuropathology.

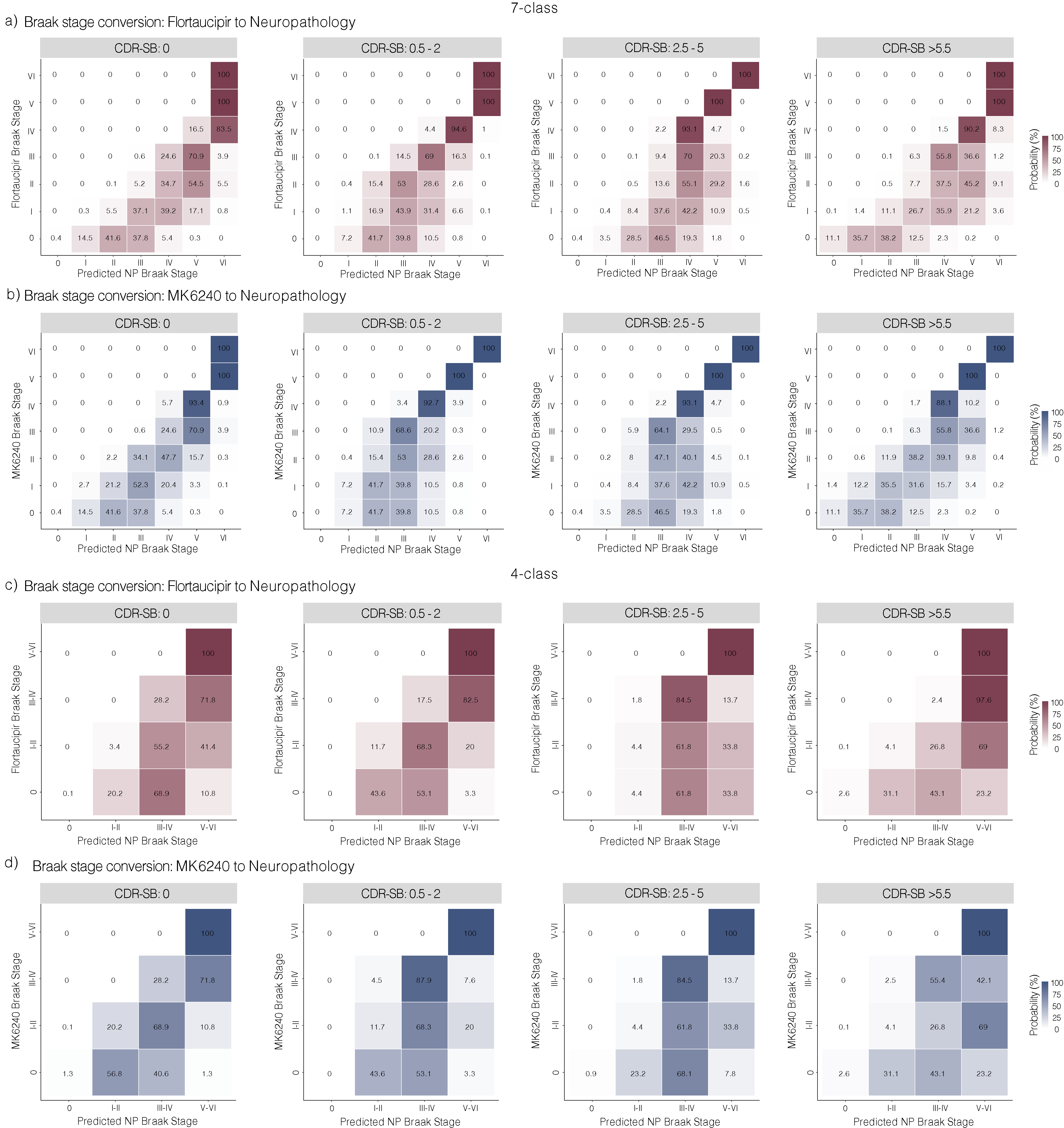

**Supplementary Figure 12. Tau-PET to neuropathological Braak stage conversion probabilities.**

Probability-based conversion matrices for Braak stage conversion from Flortaucipir (**a**, **c**) and MK6240 (**b**, **d**) to neuropathological Braak stages under the 7-class (**a**, **b**) and 4-class (**c**, **d**) staging schemes. Each cell represents the probability (%) that a given tau-PET Braak stage corresponds to a given neuropathological Braak stage within a CDR-SB severity stratum, with rows summing to 100%. Probabilities were derived by matching cumulative frequencies across tau-PET and neuropathological Braak stage distributions within each CDR-SB group and were used to generate stage-conversion tables. Neuropathological Braak stage distributions were derived from the NACC cohort, whereas tau-PET Braak stage distributions were derived from the HEAD cohort. Analyses were restricted to Aβ+ individuals for tau-PET data (CERAD 2/3 for neuropathology or Centiloid ≥ 10 for PET) and excluded Braak non-conformant cases.

**Supplementary Table 1. Demographics and key characteristics of HEAD Study young participants**

| **Characteristics** | **Young (N = 38)** |
| --- | --- |
| **Age**, mean (S.D.) | 23 (± 2.0) |
| **Sex**, n (% female) | 23 (61 %) |
| **APOE *ε4***, n (% carriers) | 5 (13 %) |
| **Education**, years (S.D.) | 16 (± 2.0) |
| **CDR Global**, mean (S.D.) | 0 (± 0) |
| **CDR-SB**, mean (S.D.) | 0 (± 0) |
| **MMSE**, mean (S.D.) | 29 (± 1.0) |
| **MoCA**, mean (S.D.) | 28 (± 2.0) |
| **Aβ Status**, n (%Aβ+) | 0 (0 %) |

Missing APOE ***ε4***, N (%) = 14 (36.8%). Missing Education, N (%) = 2 (5.3%). Missing CDR, N (%) = 4 (10.5%). Missing MMSE, N (%) = 5 (13.2%). Missing MoCA, N (%) = 2 (5.3%). APOE *ɛ4*: apolipoprotein *ɛ4*; CDR = Clinical Dementia Rating; CDR-SB = Clinical Dementia Rating Sum of Boxes; MMSE = Mini-Mental State Examination; MoCA = Montreal Cognitive Assessment.

**Supplementary Table 2. Demographics and key characteristics of NACC cohort participants.**

| **Characteristics** | **CU (N = 514)** | **MCI (N = 546)** | **Dementia (N = 3362)** |
| --- | --- | --- | --- |
| **Age**, mean (S.D.) | 86 (± 9.1) ^c^ | 86 (± 10) ^c^ | 78 (± 12) ^a,b^ |
| **Sex**, n (% female) | 280 (54 %) ^b,c^ | 258 (47 %) ^a^ | 1463 (44 %) ^a^ |
| **Education**, years (S.D.) | 16 (± 5.9) ^c^ | 17 (± 9.2) | 16 (± 9.8) ^a^ |
| **CDR Global**, mean (S.D.) | 0 (± 0) | 0.50 (± 0) | 2.4 (± 0.76) |
| **CDR-SB**, mean (S.D.) | 0.041 (± 0.16) ^b,c^ | 2.1 (± 1.5) ^a,c^ | 14 (± 4.4) ^a,b^ |
| **Aβ Status**, n (%Aβ+) | 142 (28 %) ^b,c^ | 236 (43 %) ^a,c^ | 2364 (70 %)^a,b^ |
| **Time to death**, months, mean (S.D.) | 6.6 (± 3.5) ^c^ | 6.6 (± 3.6) ^c^ | 5.8 (± 3.8) ^a,b^ |
| **PMI**, hours, mean (S.D.) | 8.6 (± 21) | 7.5 (± 21) | 8.5 (± 21) |

Categorical variables were compared using χ² or Fisher’s exact tests, and continuous variables using Kruskal-Wallis tests. When overall tests were significant, pairwise comparisons were performed using Fisher’s exact tests (categorical variables) or Wilcoxon rank-sum tests (continuous variables), with false discovery rate correction. Superscript letters indicate significant pairwise differences after FDR correction. ^a^ significantly different from CU; ^b^ significantly different from MCI; ^c^ significantly different from Dementia. CDR = Clinical Dementia Rating; CDR-SB = Clinical Dementia Rating Sum of Boxes; Time to death = interval between the last CDR assessment and death; PMI = Postmortem interval, i.e., time between death and brain collection and fixation.

**Supplementary Table 3. Tau PET Braak Stage stratification by clinical group.**

| **7-class Braak staging** | | | | | | |
| --- | --- | --- | --- | --- | --- | --- |
| **Group** | CU Aβ- (N=253) | | MCI Aβ- (N=75) | | Dementia Aβ- (N=4) | |
| **Braak Stage** | **Flortaucipir** | **MK6240** | **Flortaucipir** | **MK6240** | **Flortaucipir** | **MK6240** |
| Braak 0 | 190 (75 %) | 167 (66 %) | 46 (61 %) | 41 (55 %) | 2 (50 %) | 2 (50 %) |
| Braak I | 34 (13 %) | 25 (10 %) | 12 (16 %) | 11 (15 %) | 0 (0 %) | 0 (0 %) |
| Braak II | 7 (3 %) | 16 (6 %) | 6 (8 %) | 5 (7 %) | 1 (25 %) | 0 (0 %) |
| Braak III | 1 (0 %) | 9 (4 %) | 1 (1 %) | 4 (5 %) | 0 (0 %) | 1 (25 %) |
| Braak IV | 4 (2 %) | 9 (4 %) | 2 (3 %) | 6 (8 %) | 0 (0 %) | 0 (0 %) |
| Braak V | 1 (0 %) | 0 (0 %) | 0 (0 %) | 0 (0 %) | 0 (0 %) | 0 (0 %) |
| Braak VI | 0 (0 %) | 6 (2 %) | 0 (0 %) | 0 (0 %) | 0 (0 %) | 0 (0 %) |
| Non-conformants | 16 (6 %) | 21 (8 %) | 8 (11 %) | 8 (11 %) | 1 (25 %) | 1 (25 %) |
| **Group** | CU Aβ+ (N=109) | | MCI Aβ+ (N=150) | | Dementia Aβ+ (N=53) | |
| **Braak Stage** | **Flortaucipir** | **MK6240** | **Flortaucipir** | **MK6240** | **Flortaucipir** | **MK6240** |
| Braak 0 | 52 (48 %) | 47 (43 %) | 21 (14 %) | 13 (9 %) | 6 (11 %) | 3 (6 %) |
| Braak I | 27 (25 %)* | 11 (10 %)* | 14 (9 %)* | 3 (2 %)* | 2 (4 %) | 0 (0 %) |
| Braak II | 10 (9 %) | 19 (17 %) | 21 (14 %) | 22 (15 %) | 2 (4 %) | 1 (2 %) |
| Braak III | 2 (2 %) | 9 (8 %) | 10 (7 %) | 14 (9 %) | 1 (2 %) | 2 (4 %) |
| Braak IV | 3 (3 %) | 5 (5 %) | 31 (21 %) | 35 (23 %) | 8 (15 %) | 3 (6 %) |
| Braak V | 0 (0 %) | 2 (2 %) | 21 (14 %) | 21 (14 %) | 17 (32 %) | 8 (15 %) |
| Braak VI | 0 (0 %) | 2 (2 %) | 15 (10 %)* | 34 (23 %)* | 14 (26 %)* | 32 (60 %)** |
| Non-conformants | 15 (14 %) | 14 (13 %) | 17 (11 %) | 8 (5 %) | 3 (6 %) | 4 (8 %) |
| **4-class Braak staging** | | | | | | |
| **Group** | CU Aβ- (N=253) | | MCI Aβ- (N=75) | | Dementia Aβ- (N=4) | |
| **Braak Stage** | **Flortaucipir** | **MK6240** | **Flortaucipir** | **MK6240** | **Flortaucipir** | **MK6240** |
| Braak 0 | 230 (91 %) | 194 (77 %) | 60 (80 %) | 52 (69 %) | 2 (50 %) | 2 (50 %) |
| Braak I–II | 10 (4 %) | 31 (12 %) | 7 (9 %) | 9 (12 %) | 1 (25 %) | 0 (0 %) |
| Braak III–IV | 7 (3 %) | 13 (5 %) | 4 (5 %) | 10 (13 %) | 0 (0 %) | 1 (25 %) |
| Braak V–VI | 2 (1 %) | 6 (2 %) | 0 (0 %) | 0 (0 %) | 0 (0 %) | 0 (0 %) |
| Non-conformants | 4 (2 %) | 9 (4 %) | 4 (5 %) | 4 (5 %) | 1 (25 %) | 1 (25 %) |
| **Group** | CU Aβ+ (N=109) | | MCI Aβ+ (N=150) | | Dementia Aβ+ (N=53) | |
| **Braak Stage** | **Flortaucipir** | **MK6240** | **Flortaucipir** | **MK6240** | **Flortaucipir** | **MK6240** |
| Braak 0 | 79 (72 %)* | 60 (55 %)* | 36 (24 %)* | 16 (11 %)* | 8 (15 %) | 4 (8 %) |
| Braak I–II | 18 (17 %) | 28 (26 %) | 32 (21 %) | 30 (20 %) | 2 (4 %) | 3 (6 %) |
| Braak III–IV | 5 (5 %) | 11 (10 %) | 44 (29 %) | 48 (32 %) | 11 (21 %) | 6 (11 %) |
| Braak V–VI | 0 (0 %) | 6 (6 %) | 31 (21 %)* | 53 (35 %)* | 32 (60 %) | 39 (74 %) |
| Non-conformants | 7 (6 %) | 4 (4 %) | 7 (5 %) | 3 (2 %) | 0 (0 %) | 1 (2 %) |

Differences in Braak stage frequency between Flortaucipir and MK6240 were assessed using stage-specific χ² tests or Fisher's exact tests, as appropriate, comparing the proportion of individuals classified in each Braak stage by each tracer within each clinical-Aβ group. One test was performed per Braak stage within each clinical-Aβ stratum, and false discovery rate (FDR) correction was applied to account for multiple comparisons. * FDR-adjusted p < 0.05; ** FDR-adjusted p < 0.01.

**Supplementary Table 4. 7-class - Quantitative summary of regional differences in abnormal voxel involvement between MK6240 and Flortaucipir..**

| FTP Braak Stage | ROI | Mean Δ abnormal voxels (%) (95% CI) | Participants with  MK6240 > Flortaucipir (%) |
| --- | --- | --- | --- |
| 0 | I | 9.2 (3.6 to 14.8) | 23.2 |
|  | II | 4.3 (1.4 to 7.1) | 33.3 |
|  | III | 2.8 (0.5 to 5.1) | 49.3 |
|  | IV | 0.1 (-1.1 to 1.2) | 44.9 |
|  | V | -0.2 (-0.9 to 0.5) | 49.3 |
|  | VI | -0.5 (-1.4 to 0.3) | 40.6 |
| I | I | 33.0 (23.1 to 43.0) | 76.9 |
|  | II | 21.1 (15.8 to 26.3) | 82.1 |
|  | III | 9.5 (5.4 to 13.6) | 79.5 |
|  | IV | 4.9 (1.7 to 8.2) | 61.5 |
|  | V | 2.9 (0.6 to 5.2) | 61.5 |
|  | VI | 1.6 (-0.4 to 3.5) | 51.3 |
| II | I | 19.3 (13.8 to 24.9) | 96.9 |
|  | II | 13.1 (9.9 to 16.3) | 90.6 |
|  | III | 15.6 (10.9 to 20.4) | 93.8 |
|  | IV | 8.2 (4.2 to 12.3) | 71.9 |
|  | V | 3.2 (1.0 to 5.3) | 62.5 |
|  | VI | 2.1 (0.1 to 4.0) | 59.4 |
| III | I | 7.4 (2.3 to 12.5) | 81.8 |
|  | II | 6.2 (2.6 to 9.8) | 90.9 |
|  | III | 14.6 (6.8 to 22.4) | 90.9 |
|  | IV | 8.0 (1.4 to 14.7) | 72.7 |
|  | V | 6.5 (1.0 to 11.9) | 72.7 |
|  | VI | 5.6 (0.3 to 10.9) | 63.6 |
| IV | I | 4.7 (1.3 to 8.1) | 69.0 |
|  | II | 4.9 (2.7 to 7.0) | 78.6 |
|  | III | 10.6 (6.9 to 14.3) | 88.1 |
|  | IV | 10.7 (7.3 to 14.1) | 88.1 |
|  | V | 10.6 (7.1 to 14.2) | 85.7 |
|  | VI | 9.4 (6.0 to 12.7) | 83.3 |
| V | I | 1.4 (-0.2 to 3.1) | 55.6 |
|  | II | 1.6 (0.4 to 2.8) | 72.2 |
|  | III | 9.4 (7.1 to 11.7) | 97.2 |
|  | IV | 8.7 (6.0 to 11.4) | 94.4 |
|  | V | 8.6 (5.4 to 11.9) | 88.9 |
|  | VI | 14.3 (10.0 to 18.6) | 88.9 |
| VI | I | 3.1 (0.9 to 5.3) | 69.0 |
|  | II | 2.6 (1.1 to 4.1) | 86.2 |
|  | III | 7.1 (4.9 to 9.4) | 96.6 |
|  | IV | 7.4 (5.2 to 9.6) | 100.0 |
|  | V | 6.9 (4.8 to 9.0) | 93.1 |
|  | VI | 12.6 (8.7 to 16.5) | 93.1 |

Mean differences in the percentage of abnormal voxels (Δ = MK6240 − Flortaucipir) are shown for each Braak ROI stratified by Flortaucipir-defined Braak stage. Values are presented as mean difference (95% confidence interval). The final column indicates the percentage of participants in whom MK6240 detected a greater proportion of abnormal voxels than Flortaucipir within each Braak region. Positive values indicate greater abnormal voxel involvement detected by MK6240.

**Supplementary Table 5. 4-class - Quantitative summary of regional differences in abnormal voxel involvement between MK6240 and Flortaucipir..**

| FTP Braak Stage | ROI | Mean Δ abnormal voxels (%) (95% CI) | Participants with  MK6240 > Flortaucipir (%) |
| --- | --- | --- | --- |
| 0 | I–II | 11.3 (8.2–14.4) | 55.6 |
|  | III–IV | 2.9 (1.3–4.6) | 56.4 |
|  | V–VI | 0.8 (-0.3–1.9) | 51.3 |
| I–II | I–II | 13.4 (9.7–17.1) | 88.5 |
|  | III–IV | 8.8 (5.6–12.0) | 75.0 |
|  | V–VI | 2.9 (1.0–4.8) | 63.5 |
| III–IV | I–II | 5.2 (3.2–7.1) | 81.7 |
|  | III–IV | 10.1 (7.0–13.1) | 81.7 |
|  | V–VI | 9.7 (6.7–12.8) | 81.7 |
| V–VI | I–II | 2.3 (1.2–3.5) | 80.6 |
|  | III–IV | 7.7 (6.2–9.2) | 96.8 |
|  | V–VI | 8.3 (6.2–10.4) | 90.3 |

Mean differences in the percentage of abnormal voxels (Δ = MK6240 − Flortaucipir) are shown for each Braak ROI stratified by Flortaucipir-defined Braak stage. Values are presented as mean difference (95% confidence interval). The final column indicates the percentage of participants in whom MK6240 detected a greater proportion of abnormal voxels than Flortaucipir within each Braak region. Positive values indicate greater abnormal voxel involvement detected by MK6240.

**Supplementary Table 6. 7-class -Biomarker and clinical differences in MK6240-upstaged individuals**

| Variable | **Braak Stage** | **Concordant**  **mean (S.D.)** | **MK-upstaged**  **mean (S.D.)** | **Cohens d** | **T-value** | **df** | **p value** |
| --- | --- | --- | --- | --- | --- | --- | --- |
| **Centiloid** | 0 | 39.2 ± 36.9 | 64.4 ± 42.3 | 0.66 | -2.14 | 22.34 | 0.04* |
|  | I | 25.0 ± 16.9 | 64.4 ± 41.0 | 1.04 | -3.93 | 24.71 | 0.0006*** |
|  | II | 80.0 ± 56.0 | 81.8 ± 29.0 | 0.04 | -0.10 | 12.89 | 0.93 |
|  | III | 80.0 ± 41.3 | 80.0 ± 25.8 | 0.00 | 0.00 | 6.71 | 0.99 |
|  | IV | 108.0 ± 34.6 | 117.1 ± 39.4 | 0.24 | -0.75 | 32.78 | 0.46 |
|  | V | 116.6 ± 28.9 | 116.4 ± 38.6 | -0.01 | 0.01 | 30.25 | 0.99 |
| **CDR-SB** | 0 | 0.7 ± 1.4 | 1.2 ± 1.6 | 0.34 | -1.15 | 23.14 | 0.26 |
|  | I | 0.5 ± 1.3 | 1.3 ± 1.7 | 0.48 | -1.32 | 11.48 | 0.21 |
|  | II | 2.1 ± 2.3 | 1.2 ± 1.4 | -0.49 | 1.13 | 13.72 | 0.28 |
|  | III | 1 ± 1 | 1.9 ± 1.5 | 0.70 | -1.11 | 6.93 | 0.3 |
|  | IV | 2.2 ± 1.2 | 3.9 ± 2.2 | 0.93 | -3.14 | 35.47 | 0.003** |
|  | V | 3.7 ± 1.7 | 5.1 ± 3.1 | 0.53 | -1.67 | 30.21 | 0.11 |
| **MoCA** | 0 | 25.9 ± 3.4 | 24.7 ± 4.5 | -0.35 | 1.04 | 20.44 | 0.31 |
|  | I | 25.6 ± 4.1 | 24.4 ± 3.9 | -0.31 | 0.71 | 8.88 | 0.49 |
|  | II | 25.6 ± 3.7 | 24.2 ± 3.9 | -0.37 | 0.99 | 21.21 | 0.33 |
|  | III | 23.8 ± 5.0 | 22.4 ± 3.4 | -0.33 | 0.52 | 7.11 | 0.62 |
|  | IV | 21.6 ± 4.8 | 17.8 ± 5.0 | -0.77 | 2.34 | 30.93 | 0.026* |
|  | V | 18.2 ± 5.4 | 17.3 ± 5.8 | -0.15 | 0.40 | 27.04 | 0.69 |
| **MMSE** | 0 | 28.6 ± 1.6 | 27.3 ± 3.2 | -0.62 | 1.56 | 17.46 | 0.14 |
|  | I | 27.7 ± 2.6 | 27.4 ± 2.8 | -0.11 | 0.26 | 9.78 | 0.8 |
|  | II | 28.3 ± 3.2 | 26.6 ± 2.5 | -0.62 | 1.53 | 16.47 | 0.15 |
|  | III | 25.8 ± 3.8 | 26.4 ± 1.5 | 0.21 | -0.33 | 5.22 | 0.76 |
|  | IV | 25.6 ± 3.2 | 23.0 ± 4.8 | -0.61 | 1.98 | 35.94 | 0.056 |
|  | V | 21.7 ± 5.3 | 21.8 ± 6.1 | 0.02 | -0.05 | 28.34 | 0.96 |
| **p-tau217** | 0 | 0.4 ± 0.2 | 0.7 ± 0.3 | 1.54 | -3.08 | 6.28 | 0.02* |
|  | I | 0.5 ± 0.2 | 0.8 ± 0.3 | 1.01 | -2.92 | 13.45 | 0.012* |
|  | II | 0.6 ± 0.3 | 0.9 ± 0.3 | 0.76 | -1.40 | 7.56 | 0.2 |
|  | III | 0.6 ± 0.3 | 0.9 ± 0.2 | 1.05 | -1.13 | 2.12 | 0.37 |
|  | IV | 1.1 ± 0.3 | 1.3 ± 0.6 | 0.41 | -0.96 | 16.62 | 0.35 |
|  | V | 1.4 ± 0.6 | 1.1 ± 0.3 | -0.67 | 1.56 | 16.41 | 0.14 |
| **GFAP** | 0 | 97.4 ± 37.2 | 163.5 ± 61.4 | 1.61 | -2.56 | 5.65 | 0.045* |
|  | I | 134.0 ± 40.3 | 149.6 ± 66.2 | 0.25 | -0.66 | 10.54 | 0.52 |
|  | II | 125.5 ± 67.5 | 142.2 ± 55.3 | 0.28 | -0.43 | 4.89 | 0.68 |
|  | III | 190.4 ± 82.2 | 85.9 ± 24.8 | -2.03 | 1.75 | 1.12 | 0.31 |
|  | IV | 155.7 ± 56.6 | 184.0 ± 66.2 | 0.45 | -1.00 | 16.45 | 0.33 |
|  | V | 199.0 ± 93.1 | 206.4 ± 73.5 | 0.09 | -0.19 | 16.83 | 0.85 |

Comparison of biological and clinical measures between individuals assigned to the same Braak stage by Flortaucipir and MK6240 (Concordant) and those assigned to a higher Braak stage by MK6240 (MK6240-upstaged), stratified by Flortaucipir-defined Braak stage under the 7-class Braak staging scheme. Individuals classified as Braak non-conformant and those assigned to a higher Braak stage by Flortaucipir than MK6240 were excluded from the analysis. Continuous variables are presented as mean ± standard deviation (S.D.) and were compared using Welch two-sample t-tests. Analyses were restricted to Aβ-positive individuals (Centiloid ≥10).

**Supplementary Table 7. 4-class -Biomarker and clinical differences in MK6240-upstaged individuals**

| Variable | **Braak Stage** | **Concordant**  **mean (S.D.)** | **MK-upstaged**  **mean (S.D.)** | **Cohens d** | **T-value** | **df** | **p value** |
| --- | --- | --- | --- | --- | --- | --- | --- |
| **Centiloid** | 0 | 38.5 ± 34.2 | 64.3 ± 40.0 | 0.71 | -3.62 | 85.48 | 0.0005*** |
|  | I - II | 78.3 ± 48.1 | 82.2 ± 31.2 | 0.1 | -0.34 | 41.38 | 0.74 |
|  | III - IV | 104.7 ± 32.6 | 114.9 ± 38.0 | 0.29 | -1.05 | 50.32 | 0.29 |
|  | V - VI | 0.6 ± 1.4 | 1.2 ± 1.6 | 0.43 | -2.18 | 86.44 | 0.032* |
| **CDR-SB** | 0 | 1.4 ± 1.9 | 1.5 ± 1.3 | 0.01 | -0.04 | 42.68 | 0.97 |
|  | I - II | 2.8 ± 2.5 | 3.8 ± 2.0 | 0.43 | -1.5 | 41.09 | 0.14 |
|  | III - IV | 26.1 ± 3.7 | 23.5 ± 4.9 | -0.62 | 3.07 | 77.44 | 0.003** |
|  | V - VI | 25.1 ± 3.2 | 23.8 ± 3.9 | -0.39 | 1.34 | 44.96 | 0.19 |
| **MoCA** | 0 | 20.6 ± 5.6 | 18.7 ± 4.6 | -0.37 | 1.32 | 42.32 | 0.20 |
|  | I - II | 28.5 ± 1.9 | 26.7 ± 3.1 | -0.75 | 3.59 | 67.53 | 0.0006*** |
|  | III - IV | 27.4 ± 2.7 | 26.5 ± 2.7 | -0.32 | 1.13 | 46.93 | 0.26 |
|  | V - VI | 25.0 ± 4.1 | 23.6 ± 4.4 | -0.32 | 1.18 | 48.79 | 0.24 |
| **MMSE** | 0 | 0.5 ± 0.3 | 0.8 ± 0.4 | 0.95 | -4.49 | 69.88 | 0.00003*** |
|  | I - II | 0.7 ± 0.2 | 1.1 ± 0.4 | 1.01 | -3.42 | 33.49 | 0.0017** |
|  | III - IV | 1.2 ± 0.4 | 1.3 ± 0.6 | 0.19 | -0.73 | 47.55 | 0.47 |
|  | V - VI | 112.6 ± 54.6 | 163.4 ± 83.0 | 0.76 | -3.5 | 64.21 | 0.0009*** |
| **p-tau217** | 0 | 157.8 ± 62.8 | 188.3 ± 83.7 | 0.41 | -1.41 | 40.8 | 0.17 |
|  | I - II | 215.0 ± 110.1 | 204.6 ± 78.2 | -0.11 | 0.38 | 37.99 | 0.70 |
|  | III - IV | 38.5 ± 34.2 | 64.3 ± 40.0 | 0.71 | -3.62 | 85.48 | 0.0005*** |
|  | V - VI | 78.3 ± 48.1 | 82.2 ± 31.2 | 0.1 | -0.34 | 41.38 | 0.74 |
| **GFAP** | 0 | 104.7 ± 32.6 | 114.9 ± 38.0 | 0.29 | -1.05 | 50.32 | 0.30 |
|  | I - II | 0.6 ± 1.4 | 1.2 ± 1.6 | 0.43 | -2.18 | 86.44 | 0.032* |
|  | III - IV | 1.4 ± 1.9 | 1.5 ± 1.3 | 0.01 | -0.04 | 42.68 | 0.97 |
|  | V - VI | 2.8 ± 2.5 | 3.8 ± 2.0 | 0.43 | -1.5 | 41.09 | 0.14 |

Comparison of biological and clinical measures between individuals assigned to the same Braak stage by Flortaucipir and MK6240 (Concordant) and those assigned to a higher Braak stage by MK6240 (MK6240-upstaged), stratified by Flortaucipir-defined Braak stage under the 4-class Braak staging scheme. Individuals classified as Braak non-conformant and those assigned to a higher Braak stage by Flortaucipir than MK6240 were excluded from the analysis. Continuous variables are presented as mean ± standard deviation (S.D.) and were compared using Welch two-sample t-tests. Analyses were restricted to Aβ-positive individuals (Centiloid ≥10).

**Supplementary Table 8. Differences between neuropathology and MK6240 PET positivity across CDR-SB groups.**

| 7-class Braak staging | | | | | | |
| --- | --- | --- | --- | --- | --- | --- |
| **CDR-SB** | **Stage** | **% Neuropathology** | **%**  **MK6240** | **Odds Ratio** | **P value** | **P value**  **FRD adjusted** |
| 0 | I | 98.4 | 53.2 | 0.02 | p < 0.0001 | p < 0.0001**** |
|  | II | 86.8 | 41.8 | 0.11 | p < 0.0001 | p < 0.0001**** |
|  | III | 66.7 | 19 | 0.12 | p < 0.0001 | p < 0.0001**** |
|  | IV | 36.4 | 8.9 | 0.17 | p < 0.0001 | p < 0.0001**** |
|  | V | 17.1 | 3.8 | 0.19 | p < 0.01 | 0.013* |
|  | VI | 3.1 | 2.5 | 0.81 | 1 | 1 |
| 0.5 - 2 | I | 100 | 86.4 | 0.02 | p < 0.0001 | p < 0.001*** |
|  | II | 95.9 | 86.4 | 0.28 | 0.03 | 0.06 |
|  | III | 81.4 | 71.2 | 0.57 | 0.11 | 0.19 |
|  | IV | 58.6 | 54.2 | 0.84 | 0.57 | 0.68 |
|  | V | 31.7 | 27.1 | 0.8 | 0.52 | 0.65 |
|  | VI | 6.2 | 23.7 | 4.7 | p < 0.001 | p < 0.01** |
| 2.5 - 5 | I | 99.5 | 94.4 | 0.09 | 0.02 | 0.04* |
|  | II | 98.4 | 92.2 | 0.2 | 0.02 | 0.04* |
|  | III | 92.9 | 83.3 | 0.38 | 0.02 | 0.04* |
|  | IV | 78 | 78.9 | 1.05 | 0.87 | 0.91 |
|  | V | 50.5 | 61.1 | 1.54 | 0.1 | 0.19 |
|  | VI | 18.7 | 40 | 2.9 | p < 0.001 | p < 0.001** |
| > 5.5 | I | 99.1 | 95.7 | 0.19 | 0.19 | 0.31 |
|  | II | 97.4 | 95.7 | 0.58 | 0.46 | 0.64 |
|  | III | 94.4 | 91.3 | 0.62 | 0.38 | 0.56 |
|  | IV | 90 | 87 | 0.74 | 0.5 | 0.65 |
|  | V | 80.1 | 78.3 | 0.89 | 0.8 | 0.91 |
|  | VI | 54.2 | 52.2 | 0.92 | 0.8 | 0.91 |
| 4-class Braak staging | | | | | | |
| **CDR-SB** | **Stage** | **% Neuropathology** | **%**  **MK6240** | **Odds Ratio** | **P value** | **P value**  **FRD adjusted** |
| 0 | I–II | 98.4 | 44.9 | 0.09 | p < 0.0001 | p < 0.0001**** |
|  | III–IV | 66.7 | 15.7 | 0.01 | p < 0.0001 | p < 0.0001**** |
|  | V–VI | 17.1 | 4.5 | 0.23 | p < 0.01 | p < 0.01** |
| 0.5 - 2 | I–II | 100 | 85.1 | 0.36 | p < 0.0001 | p < 0.0001**** |
|  | III–IV | 81.4 | 60.8 | 0.02 | p < 0.001 | p < 0.01** |
|  | V–VI | 31.7 | 24.3 | 0.69 | 0.26 | 0.34 |
| 2.5 - 5 | I–II | 99.5 | 92.7 | 0.29 | p < 0.01 | p < 0.01** |
|  | III–IV | 92.9 | 79.2 | 0.07 | p < 0.001 | p < 0.01** |
|  | V–VI | 50.5 | 57.3 | 1.31 | 0.28 | 0.34 |
| > 5.5 | I–II | 99.1 | 96 | 0.68 | 0.21 | 0.31 |
|  | III–IV | 94.4 | 92 | 0.21 | 0.65 | 0.65 |
|  | V–VI | 80.1 | 72 | 0.64 | 0.32 | 0.34 |

Differences in Braak stage–specific tau positivity frequency between MK6240 and neuropathology were assessed using stage-specific χ² tests or Fisher's exact tests, as appropriate, comparing the proportion of positive individuals within each CDR-SB group. One test was performed per Braak stage within each clinical group, and false discovery rate (FDR) correction was applied to account for multiple comparisons. * FDR-adjusted; p < 0.05; ** FDR-adjusted p < 0.01; *** FDR-adjusted p < 0.001; **** FDR-adjusted p < 0.0001.

**Supplementary Table 9. Differences between neuropathology and Flortaucipir PET positivity across CDR-SB groups.**

| 7-class Braak staging | | | | | | |
| --- | --- | --- | --- | --- | --- | --- |
| **CDR-SB** | **Stage** | **% Neuropathology** | **% Flortaucipir** | **Odds Ratio** | **P value** | **P value**  **FRD adjusted** |
| 0 | I | 98.4 | 45.6 | 0.01 | p < 0.0001 | p < 0.0001**** |
|  | II | 86.8 | 17.7 | 0.03 | p < 0.0001 | p < 0.0001**** |
|  | III | 66.7 | 5.1 | 0.03 | p < 0.0001 | p < 0.0001**** |
|  | IV | 36.4 | 3.8 | 0.07 | p < 0.0001 | p < 0.0001**** |
|  | V | 17.1 | 0 | 0.03 | p < 0.001 | p < 0.001** |
|  | VI | 3.1 | 0 | 0.18 | 0.3 | 0.33 |
| 0.5 - 2 | I | 100 | 81.4 | 0.01 | p < 0.0001 | p < 0.0001**** |
|  | II | 95.9 | 69.5 | 0.1 | p < 0.0001 | p < 0.0001**** |
|  | III | 81.4 | 50.8 | 0.24 | p < 0.0001 | p < 0.0001**** |
|  | IV | 58.6 | 39 | 0.45 | 0.01 | 0.02* |
|  | V | 31.7 | 20.3 | 0.55 | 0.1 | 0.14 |
|  | VI | 6.2 | 13.6 | 2.37 | 0.1 | 0.14 |
| 2.5 - 5 | I | 99.5 | 90 | 0.05 | p < 0.001 | p < 0.001** |
|  | II | 98.4 | 81.1 | 0.07 | p < 0.0001 | p < 0.0001**** |
|  | III | 92.9 | 71.1 | 0.19 | p < 0.0001 | p < 0.0001**** |
|  | IV | 78 | 67.8 | 0.59 | 0.07 | 0.11 |
|  | V | 50.5 | 43.3 | 0.75 | 0.26 | 0.3 |
|  | VI | 18.7 | 17.8 | 0.94 | 0.86 | 0.86 |
| > 5.5 | I | 99.1 | 95.7 | 0.19 | 0.19 | 0.23 |
|  | II | 97.4 | 91.3 | 0.28 | 0.12 | 0.16 |
|  | III | 94.4 | 87 | 0.4 | 0.14 | 0.18 |
|  | IV | 90 | 87 | 0.74 | 0.5 | 0.52 |
|  | V | 80.1 | 60.9 | 0.39 | 0.03 | 0.06 |
|  | VI | 54.2 | 21.7 | 0.23 | p < 0.01 | p < 0.01** |
| 4-class Braak staging | | | | | | |
| **CDR-SB** | **Stage** | **% Neuropathology** | **% Flortaucipir** | **Odds Ratio** | **P value** | **P value**  **FRD adjusted** |
| 0 | I–II | 98.4 | 23.6 | 0.03 | p < 0.0001 | p < 0.0001**** |
|  | III–IV | 66.7 | 5.6 | 0 | p < 0.0001 | p < 0.0001**** |
|  | V–VI | 17.1 | 0 | 0.03 | p < 0.0001 | p < 0.0001**** |
| 0.5 - 2 | I–II | 100 | 67.6 | 0.16 | p < 0.0001 | p < 0.0001**** |
|  | III–IV | 81.4 | 40.5 | 0.01 | p < 0.0001 | p < 0.0001**** |
|  | V–VI | 31.7 | 17.6 | 0.46 | 0.026 | 0.03* |
| 2.5 - 5 | I–II | 99.5 | 82.3 | 0.16 | p < 0.0001 | p < 0.0001**** |
|  | III–IV | 92.9 | 67.7 | 0.03 | p < 0.0001 | p < 0.0001**** |
|  | V–VI | 50.5 | 36.5 | 0.56 | 0.025 | 0.03* |
| > 5.5 | I–II^#^ | 99.1 | 92 | 0.43 | 0.023 | 0.03* |
|  | III–IV^#^ | 94.4 | 88 | 0.1 | 0.17 | 0.17 |
|  | V–VI | 80.1 | 56 | 0.32 | p < 0.01 | p < 0.01** |

Differences in Braak stage–specific tau positivity frequency between Flortaucipir and neuropathology were assessed using stage-specific χ² tests or Fisher's exact tests, as appropriate, comparing the proportion of positive individuals within each CDR-SB group. One test was performed per Braak stage within each clinical group, and false discovery rate (FDR) correction was applied to account for multiple comparisons. * FDR-adjusted; p < 0.05; ** FDR-adjusted p < 0.01; *** FDR-adjusted p < 0.001; **** FDR-adjusted p < 0.0001.

**Supplementary Table 10. Scanners and Aβ PET tracers used across HEAD Study sites**

| **Site** | **MRI scanner** | **Aβ PET scanner** | **Aβ PET tracer** | **Tau PET scanner** |
| --- | --- | --- | --- | --- |
| **Brown University** | SIEMENS Prisma Fit | SIEMENS Biograph mCT 4R PET/CT | [^18^F]Florbetaben | SIEMENS Biograph mCT 4R PET/CT |
| **Houston Methodist Research Institute** | SIEMENS Vida | SIEMENS Biograph Vision 600 Edge PET/CT;  SIEMENS Biograph Vision Quadra Edge PET/CT | [^11^C]PiB | SIEMENS Biograph Vision 600 Edge PET/CT;  SIEMENS Biograph Vision Quadra Edge PET/CT |
| **Lawrence Berkeley National Laboratory** | SIEMENS Prisma Fit; SIEMENS TrioTim | SIEMENS Biograph mCT TruePoint PET/CT | [^11^C]PiB | SIEMENS Biograph mCT TruePoint PET/CT |
| **Mayo Clinic** | SIEMENS Prisma | GE Discovery 690 PET/CT;  GE Discovery MI PET/CT;  SIEMENS Biograph Vision 600 PET/CT | [^11^C]PiB | GE Discovery 690 PET/CT;  GE Discovery MI PET/CT;  SIEMENS Biograph Vision 600 PET/CT |
| **McGill University** | SIEMENS Prisma Fit | GE Discovery 690 PET/CT;  SIEMENS ECAT HR+ (HRRT) PET | [^18^F]NAV4694 | GE Discovery 690 PET/CT;  SIEMENS ECAT HR+ (HRRT) PET |
| **University of California San Francisco** | SIEMENS Prisma Fit | SIEMENS Biograph Vision 600 PET/CT | [^18^F]Florbetaben | SIEMENS Biograph Vision 600 PET/CT |
| **University of Pittsburgh** | SIEMENS Prisma Fit | SIEMENS Biograph mCT PET/CT;  SIEMENS ECAT HR PET | [^11^C]PiB | SIEMENS Biograph mCT PET/CT |
| **Washington University in St. Louis** | SIEMENS Prisma Fit | SIEMENS Biograph mCT TruePoint PET/CT;  SIEMENS Biograph Vision 600 Edge PET/CT | [^11^C]PiB | SIEMENS Biograph mCT TruePoint PET/CT;  SIEMENS Biograph Vision 600 Edge PET/CT |
